# Immune Subtypes and Survival in Patients with Primary Glioma

**DOI:** 10.64898/2026.04.29.26351981

**Authors:** Yu Fang, Jiwoong Kim, Zachary J. Thompson, Youngchul Kim, Harshan Ravi, Asim Mazin, Carlos M. Moran−Segura, Jonathan V. Nguyen, Robert J. Macaulay, Filippo Veglia, Reid C. Thompson, Sajeel A. Chowdhary, Kathleen M. Egan, Natarajan Raghunand

## Abstract

**Background:** Gliomas are heterogeneous tumors with poor outcomes following current therapies, including immunotherapy. The tumor microenvironment (TME) is a critical determinant of therapeutic response in gliomas. We have classified the immune TME of gliomas by multiplex immunofluorescence (mIF).

**Methods:** Tissue taken at initial resection from 354 patients with newly−diagnosed glioma grades 2-4 were analyzed using three mIF panels of markers for T, B, and myeloid cells. Tumor cores were characterized by the relative abundances of: (i) 15 primary immune phenotypes, (ii) 96 secondary immune phenotypes, and, (iii) 29 biologically meaningful multi-marker immune phenotypes.

**Results:** Using unsupervised cluster analysis of WHO grade 4 gliomas we identified four subtypes α, β, γ, and δ that were internally reproducible. Immune subtype α was characterized by high abundance of antigen-presenting cells (APCs) and low levels of MHC II− monocytes. Subtype β was high in regulatory T cells and myeloid cells, but low in lymphocytes with effector functions. Subtype γ displayed high abundance of immune cell phenotypes, particularly lymphocytes with effector or helper functions. Subtype δ was low in lymphoid and myeloid immune phenotypes and APCs, with poorer outcomes. Grade 3 tumors could also be classified into α, β, γ, and δ subtypes, indicating generalizability of these immune TME subtypes across high grade gliomas.

**Conclusions:** We have identified internally reproducible criteria for classifying gliomas according to the immune microenvironment, findings that could aid our understanding of the natural progression of low- and high-grade gliomas and inform the rational application of immune-oncologic therapeutic interventions.

## Introduction

Glioblastoma (GBM) is a devastating primary brain tumor with a 5−year survival rate of 5−10% and limited treatment options^1^. Standard therapy for newly−diagnosed GBM comprises maximal safe surgical tumor resection followed by radiotherapy with concurrent and adjuvant temozolomide chemotherapy,^2^ despite which median overall survival (OS) rates remain low. An immunosuppressive tumor microenvironment has led to limited success of immune checkpoint inhibitors, tumor−infiltrating lymphocyte (TIL) therapy, and Chimeric Antigen Receptor (CAR) T-cell therapy of GBM in the adjuvant setting.^3^ Motivated by the scarcity of tumor−infiltrating lymphocytes (TILs) in post−resection GBM, neoadjuvant anti−PD−1^4^, and combined anti−PD−1, anti−CTLA−4 and anti−LAG3^5^, therapy are being investigated in newly−diagnosed GBM. Immune checkpoint inhibition has shown minimal activity in patients with recurrent GBM^6,7^, though alterations in the infiltrating immune cell landscape in GBM following PD−1 checkpoint blockade have been observed.^8^ In a study of patients who received immune checkpoint blockade (ICB) therapy, a subset of progenitor exhausted T cells were reported to correlate with longer OS in both brain metastasis and recurrent GBM patients.^9^ High levels of ERK1/2 phosphorylation in recurrent GBM have been noted to correlate with improved response to anti−PD−1 immunotherapy, associated with elevated expression of MHC class II in tumor−infiltrating myeloid cells and microglia.^10^

In a recent study, differences in immune cell composition across grades 2−4 adult^11^ and pediatric^12^ glioma have been investigated. The densities of tumor and immune cells were noted to decrease from the tumor core to the periphery, with the GBM core exhibiting the highest level of immunosuppression.^13^ The tumor immune contexture encompasses immune parameters that are associated with patient survival, although the strength and nature of the influence of specific immune cell types on survival varies by cancer across multiple studies.^14^ In the current study, we analyzed T, B, and myeloid cell infiltrates in tumor tissue taken at initial resection to gain insight into the association between overall survival (OS) and the number and composition of tumor immune infiltrates in 354 patients with newly−diagnosed glioma grades 2, 3 and 4.

## Methods and Materials

### Patient Population

Glioma patients for this investigation are participants in a multicenter epidemiologic cohort of glioma patients enrolled at medical centers throughout the Southeastern US, including Moffitt, from 2004−2015 ^15–18^ (**Table 1**). Subjects diagnosed up to March 2015 were enrolled and provided written consent for research on tumor biomarkers. Patient vital status is current to December 31, 2016 based on a search of the National Death Index.

**Table 1:**
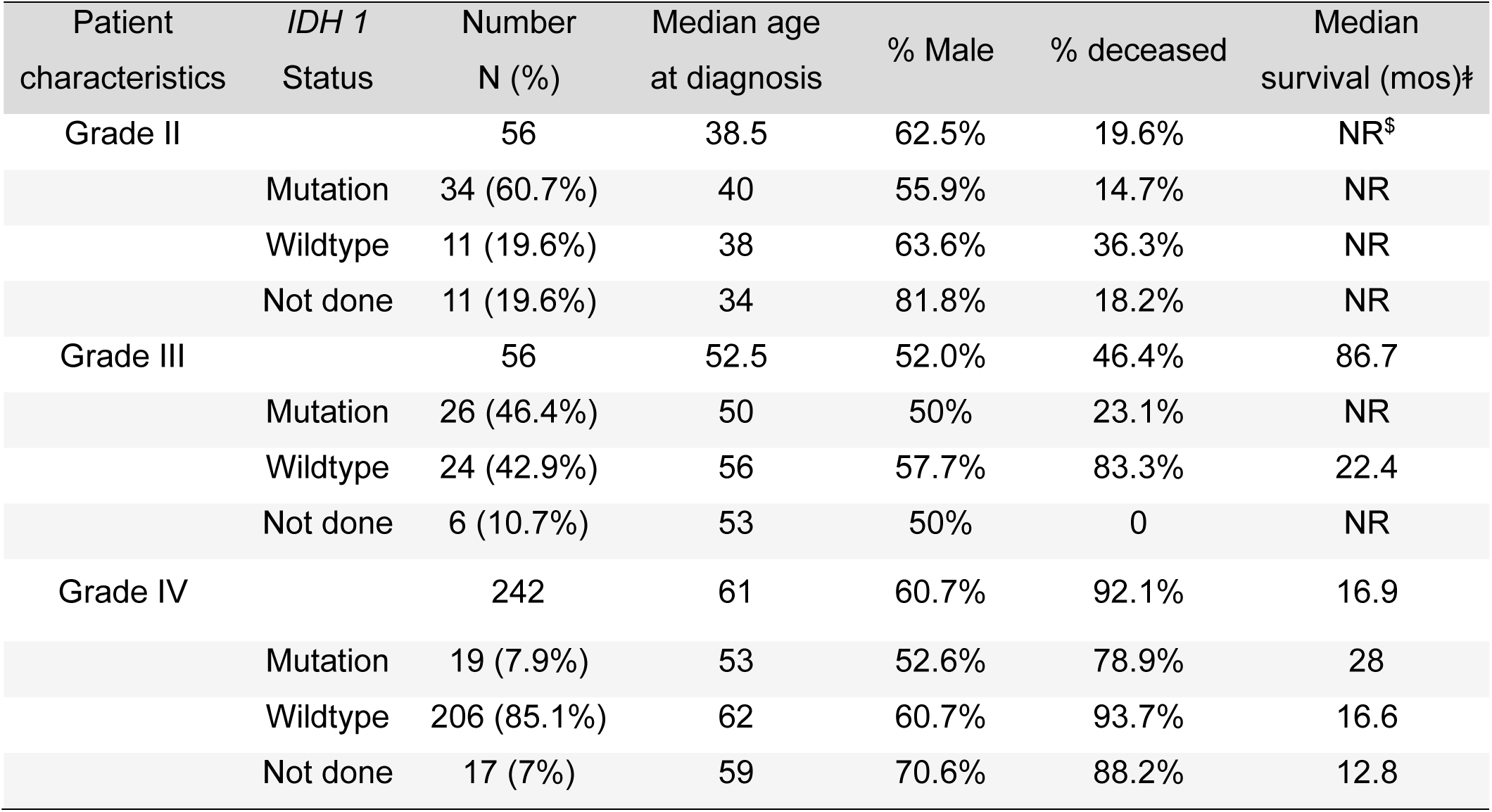
Descriptive characteristics of study participants, numbers deceased, and median survival time. Note: ꬹ Among the 45 (80.4%), 50 (89.3%) and 225 (93%) grade II, III and IV tumors, respectively, with known *IDH* mutation status. ꬸ Kaplan−Meier estimate of median time in months between tumor diagnosis and date of death or date last known alive i.e., censored. ^$^NR: Median survival not reached.

| Patient characteristics | *IDH 1* Status | Number N (%) | Median age at diagnosis | % Male | % deceased | Median survival (mos)‡ |
| --- | --- | --- | --- | --- | --- | --- |
| Grade II | | 56 | 38.5 | 62.5% | 19.6% | NR<sup>\$</sup> |
|  | Mutation | 34 (60.7%) | 40 | 55.9% | 14.7% | NR |
|  | Wildtype | 11 (19.6%) | 38 | 63.6% | 36.3% | NR |
|  | Not done | 11 (19.6%) | 34 | 81.8% | 18.2% | NR |
| Grade III |  | 56 | 52.5 | 52.0% | 46.4% | 86.7 |
|  | Mutation | 26 (46.4%) | 50 | 50% | 23.1% | NR |
|  | Wildtype | 24 (42.9%) | 56 | 57.7% | 83.3% | 22.4 |
|  | Not done | 6 (10.7%) | 53 | 50% | 0 | NR |
| Grade IV |  | 242 | 61 | 60.7% | 92.1% | 16.9 |
|  | Mutation | 19 (7.9%) | 53 | 52.6% | 78.9% | 28 |
|  | Wildtype | 206 (85.1%) | 62 | 60.7% | 93.7% | 16.6 |
|  | Not done | 17 (7%) | 59 | 70.6% | 88.2% | 12.8 |

### Construction of Tissue Microarrays and Neuropathology Review

A total of six tissue microarrays (TMAs) were constructed with specimen cores of formalin−fixed and paraffin−embedded tissue samples from 58 grade 2, 62 grade 3, and 258 grade 4 glioma tumor blocks. Across the three multiplex immunofluorescence (mIF) panels, the TMAs contained the following numbers of cores: Panel 1 included 767 cores (118 grade 2, 128 grade 3, and 521 grade 4); Panel 2 included 825 cores (129 grade 2, 145 grade 3, and 551 grade 4); and Panel 3 included 768 cores (120 grade 2, 131 grade 3, and 517 grade 4). In total, there were 56 grade 2, 56 grade 3, and 242 grade 4 glioma tumor blocks with complete measurements across all three mIF panels.

Prior to TMA construction, all cases were reviewed by a neuropathologist (RM) to confirm the diagnosis and preselect areas of tumor from which up to three 0.6 mm cores were included per tumor. Each TMA was constructed in triplicate for analysis using three mIF panels.

### Multiplex Immunofluorescence Microscopy

TMAs were immunostained using the Akoya Biosciences Opal TM 7−Color Automation IHC kit (Waltham, MA) on the Bond RX autostainer (Leica Biosystems, Vista, CA). One set of TMAs each were stained using 3 mIF panels: panel 1 with fluorescent markers of FoxP3, CD3, CD4, CD8 and PD−1, panel 2 with markers of CD11b, CD14, CD15, CD33 and MHCII, and panel 3 with markers of CD19, CD20, CD68, CD163 and CD206. All 3 panels also included DAPI to stain cell nuclei, and Glial Fibrillary Acidic Protein (GFAP) as an astrocytic marker. Autofluorescence correction was performed using negative control slides with primary and secondary antibodies but not the Opal fluorophores, DAPI, or GFAP. All slides were imaged with the Vectra3® Automated Quantitative Pathology Imaging System. Representative images of tumor cores stained using each mIF panel are shown in **Supplementary Figures 1−3**.

### IDH1 Staining

IHC of TMAs to detect mutation in *IDH1* was performed by RM using an assay that detects *IDH1* R132H mutation found in the majority of *IDH1* mutated tumors. The highly specific anti−*IDH1* R132H antibody clone H09 (Dianova) was utilized for the investigation.

### Multiplex Immunofluorescence Image Analysis

Multi−layer TIFF images were exported from InForm (Akoya) and loaded into HALO® Image Analysis Platform version 3.6.4134 (Indica Labs, New Mexico) for quantitative image analysis. Individual cells in each core on a TMA were segmented by an algorithm trained in HALO®. For each marker, a positivity threshold within the nucleus or cytoplasm was determined using a machine learning algorithm in HALO®, which was manually adjusted based on visual analysis for expected cytoplasmic, membrane or nuclear localization of each marker in a subset of slides. The entire image set was analyzed with the selected positivity thresholds. The relative location, binary positivity status per marker, and continuous fluorescence intensities per marker for each cell were exported for quantitative analysis of each core.

### Quality Control and Data processing

Tonsil tissue cores were included in each TMA for use as controls to assess staining consistency. For each marker, Z−scores were calculated within each TMA to identify potential outliers^19^. No significant staining differences across TMAs were observed (**Supplementary Table 1**).

There were a variable number of replicates per patient across the six TMAs. To generate a single patient level phenotypic value while minimizing individual differences in core selection, we summarized core level measurements using a weighted average based on total cell count in each core. Specifically, for a given phenotype, each core’s relative cell counts were weighted by its total cell counts, ensuring that cores containing more informative tissue contributed proportionally more to the final estimate.

### Construction of Secondary and Multi−marker Secondary Phenotypes

Given five immune−related markers there can be 32 (= 2^5^) possible combinations per panel, for a total of 96 possible secondary immune cell phenotypes across the three mIF panels. Considering the sparsity or even total absence of several secondary phenotypes, and based on the known biological functions of the immune markers, we grouped the 96 combinations into 29 biologically meaningful multi−marker secondary phenotypes (**Table 2**).

**Table 2:** Phenotypic features used for the identification of multi−marker secondary phenotypes. Note: Details of the secondary phenotypes included in each summed multi−marker secondary phenotype are provided in **Supplementary** Table 2. Abbreviations: −, marker not expressed; +, marker expressed; −/+, marker expressed in a subset of the cell population.

| Panel 1<br>Markers | Combined multi-marker phenotypes |  |  |  |  |  |  |  |  |  |  |  |
| --- | --- | --- | --- | --- | --- | --- | --- | --- | --- | --- | --- | --- |
|  | NK Cells | innate-like<br>T cells | double-negative<br>T cells | cytotoxic<br>T cells | T helper cells | double-positive<br>T cells | PD1<sup>+</sup>CD3<sup>-</sup><br>cells | PD1<sup>+</sup><br>T lymphocytes | FoxP3<sup>+</sup>CD3<sup>-</sup><br>cells | double-negative<br>Tregs | CD8<sup>+</sup><br>Tregs | Tregs |
| FoxP3 | – | – | – | – | – | – | – | +/– | + | + | + | + |
| CD3 | – | – | + | + | + | + | – | + | – | + | + | + |
| CD4 | – | + | – | – | + | + | +/– | +/– | +/– | – | – | + |
| CD8 | + | +/– | – | + | – | + | +/– | +/– | +/– | – | + | +/– |
| PD1 | – | – | – | – | – | – | + | + | – | – | – | – |
| Panel 2<br>Markers | Combined multi-marker phenotypes |  |  |  |  |  |  |  |  |  |  |  |
|  | CD11b<sup>-</sup>CD14<sup>-</sup><br>APCs | CD11b<sup>-</sup>CD14<sup>+</sup><br>APCs | CD11b<sup>+</sup>CD14<sup>+</sup><br>APCs | activated microglia | MHC II<sup>+</sup> neutrophils | CD14<sup>+</sup>CD11b<sup>-</sup><br>MHC II<sup>-</sup> cells | MHC II<sup>-</sup> monocytes | myeloid-like<br>cells | glial stem-like<br>cells | microglia-like<br>cells | neutrophils |  |
| CD11b | – | – | + | + | + | – | + | +/– | – | + | + |  |
| MHC II | + | + | + | + | + | – | – | – | – | – | – |  |
| CD14 | – | + | + | – | – | + | + | – | – | – | – |  |
| CD15 | +/– | +/– | +/– | – | + | +/– | +/– | +/– | + | – | + |  |
| CD33 | +/– | +/– | +/– | +/– | +/– | +/– | +/– | + | – | – | – |  |
| Panel 3<br>Markers | Combined multi-marker phenotypes |  |  |  |  |  |  |  |  |  |  |  |
|  | B cells | macrophages/microglia |  | M2 macrophages | CD206<sup>+</sup> M2-like macrophages |  | CD163<sup>+</sup> M2-like macrophages | CD68<sup>-</sup> M2-like macrophages |  |  |  |  |
| CD163 | +/– | – |  | + | – |  | + | +/– |  |  |  |  |
| CD68 | – | + |  | + | + |  | + | – |  |  |  |  |
| CD20 | +/– | +/– |  | +/– | +/– |  | – | – |  |  |  |  |
| CD19 | +/– | +/– |  | +/– | +/– |  | +/– | – |  |  |  |  |
| CD206 | +/– | – |  | + | + |  | – | +/– |  |  |  |  |

### Dimension Reduction and Cluster Analysis

To identify latent patterns within the 96−dimensional secondary phenotypes feature space across glioma grades, unsupervised clustering was performed in a fully data-driven manner, without predefined labels. Uniform Manifold Approximation and Projection (UMAP)^20^ implemented using the package “uwot” (v0.2.3)^21^ was used to generate a two−dimensional representation of the 96−dimensional secondary phenotype matrix. Prior to UMAP embedding, Principal Component analysis (PCA) was applied to the matrix to capture dominant sources of variation and reduce noise^22^. Only PCs with eigenvalues greater than 1 were retained, following the Kaiser criteria^23^. Clusters were subsequently identified in the UMAP embedding using Hierarchical Density−Based Spatial Clustering of Applications with Noise (HDBSCAN)^24^, as implemented in the package “dbscan” (v1.2.2)^25^.

To evaluate the robustness of the clustering results, the UMAP embedding was generated across 3,001 random seeds (0-3000), and HDBSCAN clustering was applied for each setting. Clustering consistency across runs and the clustering results for each grade of tumors were quantified using the Adjusted Rand Index (ARI), and the mean and standard deviation of ARI were computed^26^. To assess whether the clusters identified in grade 4 tumors generalized to other high−grade gliomas, grade 3 tumors were treated as an independent input dataset and projected onto the previously learned PCA and UMAP embeddings for grade 4, followed by cluster assignment using the HDBSCAN model trained on grade 4. Additionally, multivariable Cox proportional hazards regression analyses for OS were performed to evaluate whether the identified phenotypic clusters were associated with patient survival outcomes. Cluster membership was included as the main predictor while adjusting for age and *IDH1* status.

### Immune Subtyping of Gliomas

We explored the prevalence of primary phenotypes and multi−marker secondary phenotypes in different subtypes within a grade. The Kruskal−Wallis test was first used to identify markers showing significant differences among patient groups, especially to determine whether at least one group differed significantly from others. For markers demonstrating statistical significance in the Kruskal–Wallis test (p ≤ 0.05), post hoc pairwise comparisons were conducted using Dunn’s test^27^. False Discovery Rate (FDR)-adjusted p−values were calculated to account for multiple hypothesis testing when assessing group differences^28^. All statistical analyses were calculated using R (R version 4.4.2)and GraphPad Prism 10 (GraphPad Software, Boston, MA).

## Results

### Dataset characteristics and multi−marker secondary immune phenotypes

**Table 1** summarizes patient characteristics and survival according to glioma grade. A majority of patients were male, regardless of tumor grade. Median age at diagnosis increased with tumor grade: 38.5 years for grade 2, 52.5 years for grade 3 and 61 years for grade 4. The majority of patients with grade 4 died a median of 16.9 months after diagnosis, whereas most patients with grade 2 tumors were alive at the time of the study, with intermediate survival among patients with grade 3 tumors. *IDH1* mutation status was available for 321 (90%) of the samples: tumors were positive for *IDH1-mut* in 73.9%, 52% and 8.4% of grade 2, grade 3 and grade 4s, respectively (**Table 1**). To reduce sparsity and multicollinearity while preserving biologically interpretable tumor microenvironment states, 96 low frequency secondary phenotypes were summed into 29 biologically relevant multi−marker phenotypes (**Table 2**). The specific secondary phenotypes contributing to each multi−marker secondary phenotype are listed in **Supplementary Table 2**.

### Identification of Glioma Immune Subtypes

Within each glioma grade, PCA was first performed on the relative prevalences of the 96 secondary phenotypes, following which the Kaiser criterion^23^ was used to decide how many principal components (PCs) to include in our analysis; as summarized in **Supplementary Table 3**, 93% of the variance in grade 2 feature space, 89% in grade 3 feature space, and 86% variance in grade 4 feature space were captured by selected PCs. On these selected PCs, two−dimensional unsupervised UMAP was then performed, following which HDBSCAN was performed to identify four subtypes of grade 4 tumors (G4α, G4β, G4γ, G4δ), three subtypes of grade 3 tumors (G3ε, G3ρ, G3τ), and two subtypes of grade 2 tumors (G2φ, G2ψ) (**Figure 1A, C, E**). Kaplan−Meier analyses were performed to investigate any relationships between OS and the tumor immune subtype (**Figure 1B, D, F**).

**Figure 1.**
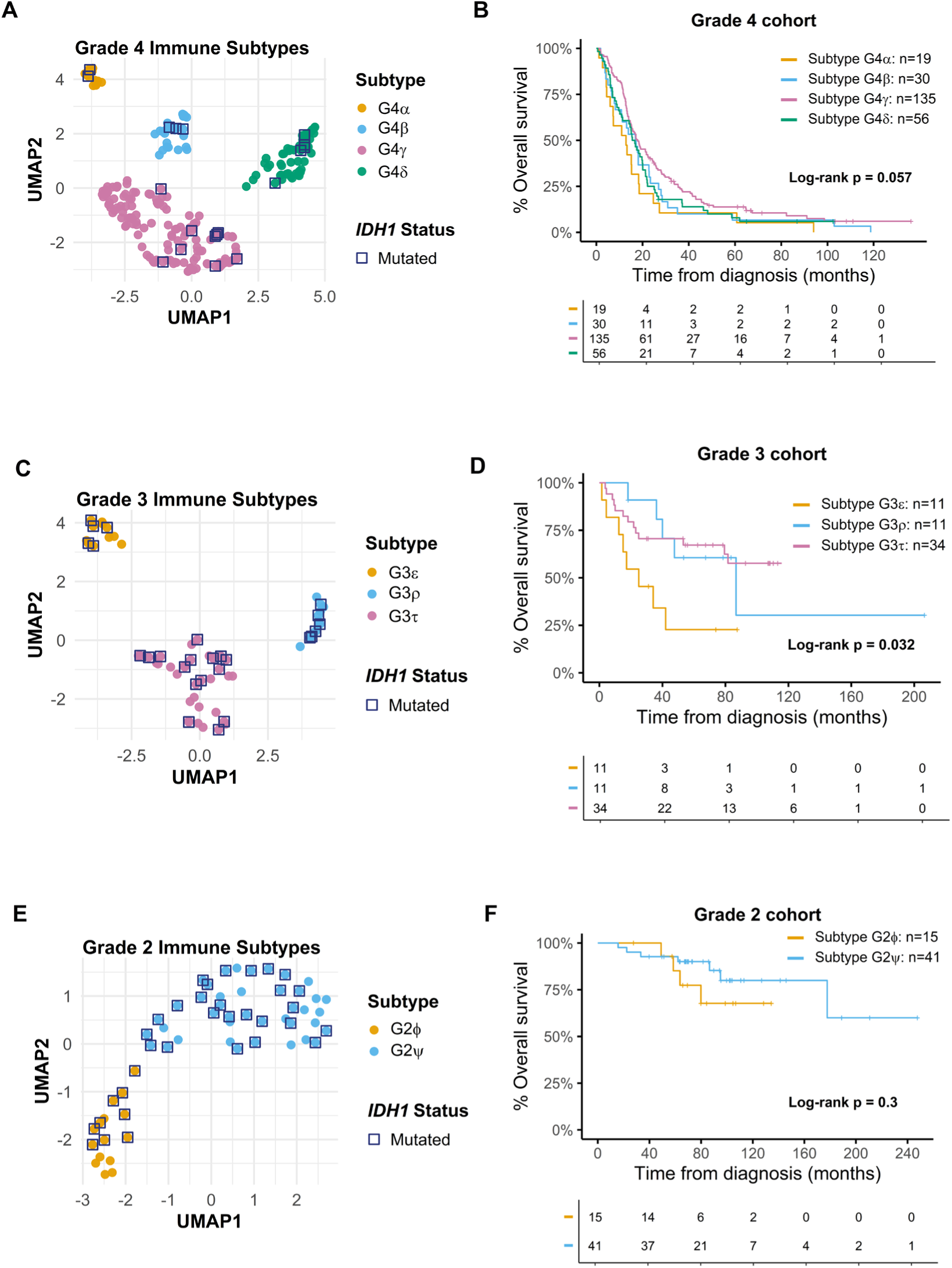
Unsupervised clustering and Kaplan−Meier plots of OS stratified by tumor subtypes. **(A)** Unsupervised UMAP projections colored by clusters identified using HDBSCAN for grade 4 tumor cohort, after removing two outliers during clustering. **(B)** Corresponding Kaplan−Meier plot of OS stratified by grade 4 tumor subtypes. **(C)** & **(E)**: Unsupervised UMAP projections colored by subtypes identified using HDBSCAN for grade 3 and grade 2 tumor cohorts. **(D)** & **(F)**: Corresponding Kaplan−Meier plots of OS stratified by tumor subtypes.

### Immune phenotyping of grade 4 glioma subtypes

As UMAP is a stochastic algorithm, we first investigated the reproducibility of our PCA−UMAP−HDBSCAN workflow for clustering grade 4 tumors. Across 3,001 random seeds to initiate UMAP embedding, and four values of minPts (the minimum number of points required to form a cluster in HDBSCAN), the mean ARI remained high (> 0.8), pointing to the stability of our clustering framework for grade 4 tumors (**Supplementary Figure 4A**). Multiple primary and multi−marker secondary phenotypes were differentially prevalent across G4α, G4β, G4γ, G4δ tumors. Violin plots of the relative abundances of all multi−marker secondary phenotypes that were significantly different between at least one pair of grade 4 tumor immune subtypes are presented in **Figure 2**, while equivalent data on primary phenotype abundances are presented in **Supplementary Figure 5**.

**Figure 2:**
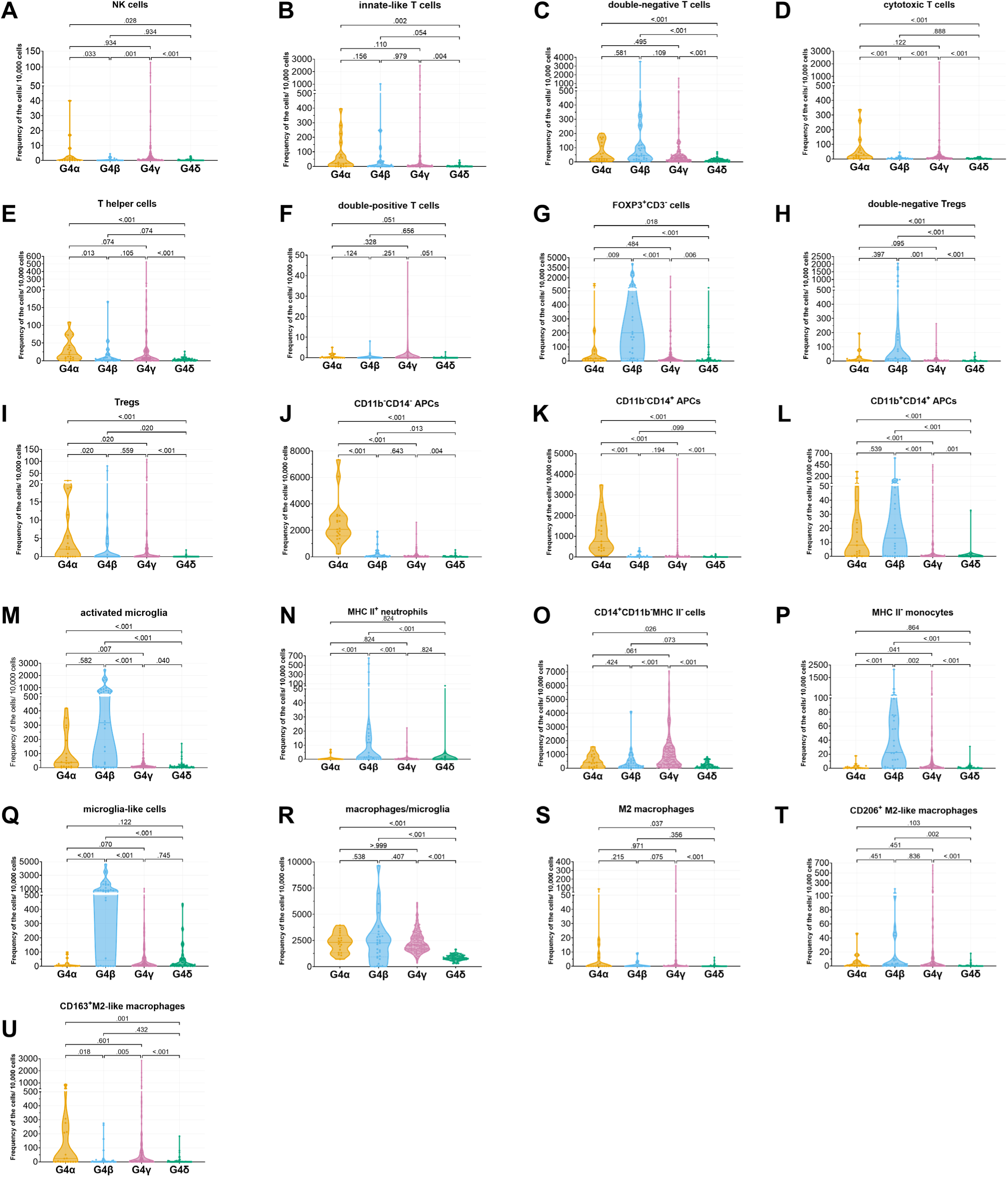
Violin plots of multi-marker secondary phenotypes significant by Kruskal−Wallis testing in grade 4 cohort, stratified by subtypes. Dunn’s post hoc tests were applied, with FDR−adjusted p−values reported.

G4α tumors had a higher abundance of the primary CD4^+^ phenotype than G4β, G4γ and G4δ tumors (**Supplementary Figure 5**). Accordingly, multiple CD4^+^ associated secondary phenotypes, including innate−like T cells, T helper cells, Tregs, and double−positive T cells, were also more abundant in G4α tumors than in G4β, G4γ and G4δ tumors (**Figure 2**). CD8^+^ immune features demonstrated a similar enrichment pattern, with primary CD8^+^ cells (**Supplementary Figure 5**), cytotoxic T cells, and double−positive T cells (**Figure 2**) all being more abundant in G4α tumors than in G4β, G4γ and G4δ tumors. The primary MHCII^+^ phenotype (**Supplementary Figure 5**), as well as five multi−marker secondary phenotypes expressing MHCII (**Figure 2**), were also relatively enriched in G4α tumors. Both CD11b^−^CD14^−^ and CD11b^−^CD14^+^ APCs were significantly enriched in G4α tumors relative to G4β, G4γ and G4δ tumors, while CD11b^+^CD14^+^ APCs tended to be higher in G4α tumors but this difference did not reach statistical significance (**Figure 2**). The primary CD163^+^ and CD68^+^ phenotypes exhibited higher levels in G4α tumors though these differences were not statistically significant (**Supplementary Figure 5**). The multi−marker secondary phenotypes macrophages/microglia, M2 macrophages, and CD163^+^ M2−like macrophages followed the same trend, namely higher abundance in G4α tumors relative to G4β, G4γ and G4δ tumors, but did not achieve statistical significance (**Figure 2**). Overall, the 19 G4α tumors were characterized by high abundance of APCs and low levels of MHC II^−^ monocytes (**Supplementary Table 4**).

G4β tumors were characterized by relative abundance of the primary FoxP3^+^ phenotype, which was significantly higher than in G4α, G4γ and G4δ tumors (**Supplementary Figure 5**). Accordingly, both FoxP3^+^ associated multi−marker secondary phenotypes, *viz.* FoxP3^+^CD3^−^ cells and double−negative Tregs, were more abundant in G4β tumors than in G4α, G4γ and G4δ tumors (**Figure 2**). Other enriched multi−marker secondary phenotypes in G4β tumors included double−negative T cells, activated microglia, MHC II^+^ neutrophils, microglia−like cells. Overall, the 30 G4β tumors were high in regulatory T cells, high in myeloid cells, including microglia and neutrophils, and low in lymphocytes with effector functions, including NK cells and cytotoxic T cells (**Supplementary Table 4**).

G4γ tumors, representing 135 patients, were the most common grade 4 immune subtype. No primary phenotype was most abundant in G4γ tumors (**Supplementary Figure 5**), though the multi−marker secondary phenotypes CD14^+^CD11b^−^MHCII^−^ cells and NK cells were more abundant in G4γ tumors than in G4α, G4β and G4δ tumors (**Figure 2**). Overall, G4γ tumors displayed high abundance of immune cell phenotypes, particularly lymphocytes with effector or helper functions, including NK cells, innate−like T cells, cytotoxic T cells, T helper cells, and double−positive T cells (**Supplementary Table 4**).

G4δ tumors, representing 56 patients, showed low abundance of lymphoid and myeloid immune phenotypes, as well as low APCs (**Figure 2**, **Supplementary Figure 5**). Specifically, G4δ tumors were characterized by low relative cell counts of NK cells, innate−like T cells, double−negative T cells, cytotoxic T cells, T helper cells, double−positive T cells, double−negative Tregs, Tregs, CD11b^−^CD14^−^ APCs, CD11b^−^CD14^+^ APCs, CD11b^+^CD14^+^ APCs, activated microglia, MHC II^−^ monocytes, and macrophages/microglia. (**Supplementary Table 4**).

We performed Kaplan−Meier curve analysis and log rank test to investigate associations between grade 4 tumor immune subtype and survival outcomes. While statistically significant differences in OS were not found, patients with G4α tumors had the shortest OS, followed by patients with G4β and G4δ tumors, while patients with G4γ tumors had the longest OS (**Figure 1B**).

### Immune phenotyping of grade 3 glioma subtypes

Within the grade 3 patient cohort, unsupervised clustering on PCA and UMAP projections of 96 secondary phenotypes identified three distinct tumor immune subtypes: G3ε, G3ρ, and G3τ (**Figure 1C**). Across 3,001 random seeds to initiate UMAP embedding, and four values of minPts (the minimum number of points required to form a cluster in HDBSCAN), the mean ARI remained high (≈ 0.8), pointing to the stability of our clustering framework for grade 3 tumors (**Supplementary Figure 4B**). Four primary phenotypes, *viz.*, FoxP3^+^, CD3^+^, CD14^+^ and CD68^+^ cells, were significantly different between at least one pair of grade 3 immune subtypes (**Supplementary Figure 6**). Correspondingly, the six multi−marker secondary immune phenotypes double−negative T cells, FoxP3^+^CD3^−^ cells, double−negative Tregs, CD8^+^ Tregs, activated microglia, and macrophages/microglia were also significantly different between at least one pair of grade 3 tumor immune subtypes (**Supplementary Figure 7**). Overall, G3ε tumors (*n* = 11) were characterized by low levels of both lymphoid and myeloid immune phenotypes, particularly double−negative T cells, FoxP3^+^CD3^−^ cells, double−negative Tregs, CD8^+^ Tregs, activated microglia and macrophage/microglia. G3ρ tumors (*n* = 11) exhibited high abundances of myeloid cells and regulatory T cells, including double−negative T cells, FoxP3^+^CD3^−^ cells, double−negative Tregs, CD8^+^ Tregs, activated microglia and macrophage/microglia. G3τ tumors (*n* = 34) were high in myeloid cells, activated microglia, and macrophage/microglia, and low in regulatory T cells, including double−negative T cells, FoxP3^+^CD3^−^ cells, double−negative Tregs and CD8^+^ Tregs (**Supplementary Figure 6**, **Supplementary Figure 7**; **Supplementary Table 5**). Patients with G3ε tumors had the shortest OS, while patients with G3ρ and G3τ tumors had comparable OS (**Figure 1D**).

### Immune phenotyping of grade 2 glioma subtypes

Within the grade 2 patient cohort, unsupervised clustering on PCA and UMAP projections of 96 secondary phenotypes identified two distinct tumor immune subtypes: G2φ and G2ψ (**Figure 1E**). Stability of our clustering framework for grade 2 tumors was moderate, with a mean ARI of 0.51-0.67 (**Supplementary Figure 4C**). Analysis of the relative abundances of immune cell phenotypes revealed that 14 multi−marker secondary phenotypes differed significantly between the two clusters, with G2ψ tumors being enriched in all of them relative to G2φ tumors (**Supplementary Figure 8**). G2ψ tumors were also enriched in nine primary immune phenotypes relative to G2φ tumors (**Supplementary Figure 9**). Overall, G2φ tumors (*n* = 15) showed uniformly lower relative abundances of both lymphoid and myeloid immune cell phenotypes relative to G2ψ tumors (*n* = 41) (**Supplementary Table 6**). Patients with G2ψ tumors tended to have longer OS relative to patients with G2φ tumors, suggesting an association between tumor enrichment in immune cell phenotypes a more favorable survival profile (**Figure 1F**).

### Mapping lower-grade glioma immune profiles onto the immune space of higher-grade gliomas

When the immune profiles of 56 grade 3 gliomas were projected onto the UMAP embedding learned on grade 4 gliomas, four immune subtypes of grade 3 tumors (G3α, G3β, G3γ, G3δ) were identifiable, with only 3 outliers (**Figure 3A**). Analogously, when the immune profiles of 56 grade 2 gliomas were into the UMAP immune space of grade 3 gliomas, three immune subtypes of grade 2 tumors (G2ε, G2ρ, and G2τ) were identifiable, with no outliers (**Figure 3C**). These results suggest that the immune TME subtypes of lower grade gliomas can be adequately described by immune TME subtype categories that were defined on the next higher grade glioma.

**Figure 3.**
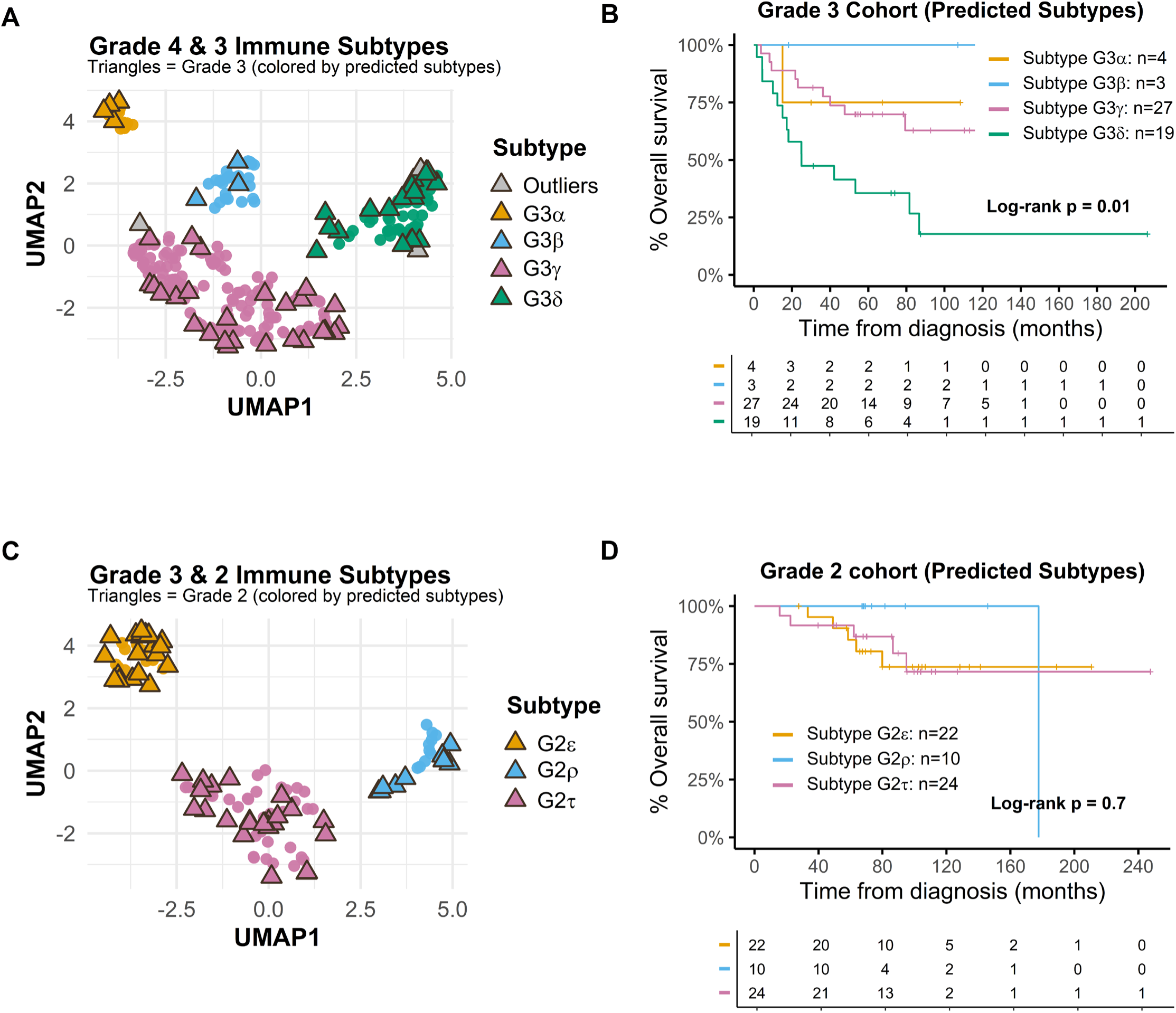
Projection of lower grade gliomas onto UMAP embedding of higher grade gliomas. **(A)** Unsupervised UMAP embedding was generated using grade 4 cohort. Grade 3 tumors were projected into the same embedding space using the trained UMAP model. Cluster assignments were predicted using the HDBSCAN model trained on grade 4 data. **(B)** Corresponding Kaplan−Meier plot of OS stratified by predicted grade 3 tumor subtypes. **(C)** Unsupervised UMAP embedding was generated using grade 3 cohort. Grade 2 tumors were projected into the same embedding space using the trained UMAP model. Cluster assignments were predicted using the HDBSCAN model trained on grade 3 data. **(D)** Corresponding Kaplan−Meier plot of OS stratified by predicted grade 2 tumor subtypes.

G3δ tumors overlapped almost completely with G3ε tumors (**Supplementary Table 7**), representing an immune subtype with low levels of both lymphoid and myeloid immune phenotypes (**Supplementary Figure 10**). G3ρ tumors overlapped largely, but not completely, with G3γ tumors (**Supplementary Table 7**) that are characterized by high abundance of immune cell phenotypes, particularly lymphocytes with effector function (**Supplementary Figure 10**). G3τ tumors also overlapped substantially with G3γ tumors (**Supplementary Table 7**). Patients with G3γ subtype had significantly better outcomes than patients with G3δ tumors (**Figure 3B**), which recapitulated outcome differences between patients with G4γ and G4δ tumors, though the latter did not achieve statistical significance (**Figure 1B**).

G2φ tumors mapped completely into the G2ε subtype (**Supplementary Table 8**), representing tumors with low levels of both lymphoid and myeloid immune phenotypes. G2ψ tumors mapped onto the immune space of all three subtypes defined on grade 3 gliomas (**Supplementary Table 8**), with a majority overlapping with the G2τ subtype that was high in myeloid cells and low in regulatory T cells. A significant fraction of G2ψ tumors also mapped into the G2ρ subtype characterized by high abundances of myeloid cells and regulatory T cells (**Supplementary Table 8**). While the τ subtype had the poorest outcome in grade 3 (**Figure 1D**), statistically significant differences in outcomes between immune subtypes were not observed in grade 2 gliomas (**Figure 3D**).

### Stratified Cox regression analysis of glioma immune subtypes

To evaluate the association between tumor immune subtypes and OS in both grade 3 and grade 4 patients, we constructed three multivariable Cox proportional hazards models (**Figure 4**). G4γ tumors, corresponding to the longest OS grade 4 subtype, were used as the reference category. Additionally, to enable reliable comparisons in multivariable Cox regression models, we excluded the small G3α (*n* = 4) and G3β (*n* = 3) tumor subtypes from all three models shown in **Figure 4**. To account for the survival differences between Grade 3 and Grade 4 patients, we used a grade-stratified Cox regression model in both grades combined model. This allowed each grade to have its own baseline hazards while estimating subtype effects.

**Figure 4.**
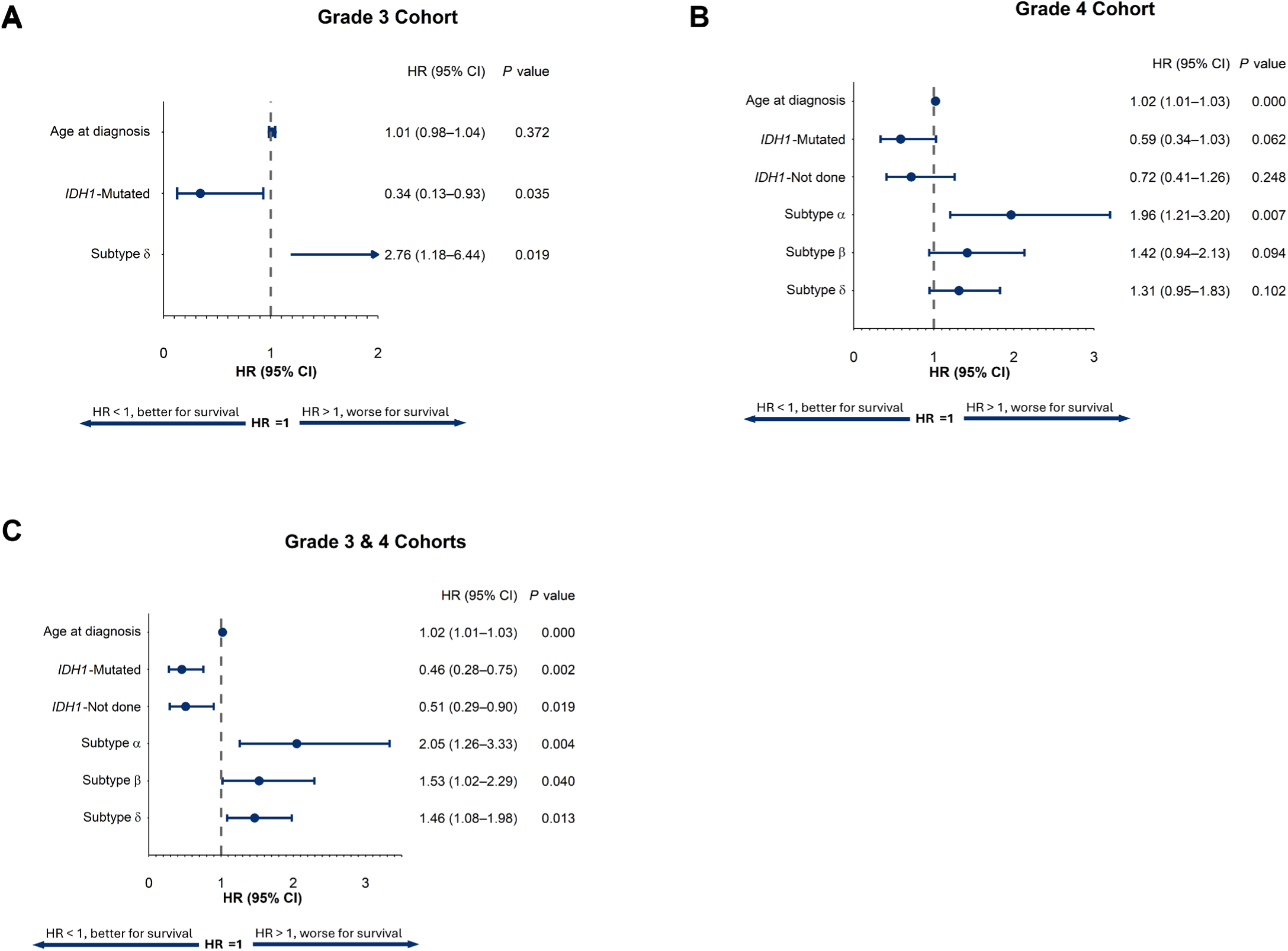
Multivariable Cox models of age, *IDH1* status and cluster results. To get more reliable results, grade 3 tumors predicted as subtype G3α (n = 4) and G3β (n = 3) were excluded from all plots in figure 4 due to the very small sample sizes. In all three models shown here, subtype γ was used as the reference group for the cluster category. **(A)** Forest plot of multivariable Cox proportional hazards models for OS, evaluating age, *IDH1* status and predicted subtypes in grade 3 cohort. Hazard ratios (HR) and 95% confidence intervals (95% CI) are shown for each variable. *IDH1*−wildtype and *IDH1* status not done were combined and used as the baseline group for comparisons with *IDH1*−mutated tumors. P values are tested by two−sided Wald test. HR > 1 indicates worse survival, while HR < 1 indicates better survival. **(B)** Forest plot of multivariable Cox proportional hazards models for OS, evaluating age, *IDH1* status and predicted clusters in grade 4 cohort. **(C)** Forest plot of multivariable Cox proportional hazards models for OS, evaluating age, *IDH1* status and predicted subtypes in grade 4 and grade 3 cohorts using a grade stratified analysis.

In grade 3 (**Figure 4A**), grade 4 (**Figure 4B**), and both grades combined (**Figure 4C**), with *IDH1-wt* as the reference category, both *IDH1-mut* and *IDH1-Not done* showed longer OS (HR < 1) relative to *IDH1-wt* tumors. In grade 4 tumors, *IDH1-mut* and *IDH1-Not done* were not significantly different compared with the baseline group, *IDH1-wt* tumors (**Figure 4B**). But in both grade 3 and the combined grade 3 and grade 4 cohort, *IDH1* status was significant (**Figure 4A; C**). Across all three models, glioma immune subtype γ (G3γ and/or G4γ) was associated with the longest OS. Immune subtype δ was associated with significantly shorter OS than immune subtype γ in both the grade 3 cohort (HR = 2.76, 95% CI: 1.18−6.44, p = 0.019) and the combined grade 3 and grade 4 analysis (HR = 1.46, 95% CI: 1.08−1.98, p = 0.013), though the trend was non-significant when grade 4 tumors were considered separately (HR = 1.31, 95% CI: 0.95−1.83, p = 0.102).

### Survival Analysis According to Relative Densities of Immune Cell Phenotypes

For the sake of completeness, we also performed multivariable Cox proportional hazards regression analyses to understand the influence of individual immune cell phenotypes, without reference to immune TME subtype, on OS in each grade. Methodological details are provided in **Supplementary Note 1**.

In patients with grade 4, higher densities of CD8^+^ Tregs (HR = 0.81, 95% CI: 0.67−0.97, p = 0.021) and MHCII^+^ neutrophils (HR = 0.88, 95% CI: 0.77−1.00, p = 0.047) were significantly associated with higher OS. Conversely, higher densities of glial stem−like cells (HR = 1.23, 95% CI: 1.07−1.42, p = 0.003), PD1^+^ cells (HR = 1.21, 95% CI: 1.05−1.41, p = 0.009), double−negative Tregs (HR = 1.22, 95% CI: 1.05−1.41, p = 0.009), and older age (HR = 1.38, 95% CI: 1.21−1.58, p < 0.001) were significantly associated with lower OS (**Supplementary Figure 11A**). Additionally, OS showed a weak negative association with the density of microglia−like cells (HR = 1.11, 95% CI: 0.98−1.27, p = 0.113). The model’s performance is illustrated by multivariable Cox regression analysis (**Supplementary Figure 11A**, C−index = 0.623, 95% CI: 0.582−0.664; **Supplementary Figure 11B**, log−rank p < 0.001), indicating a significant separation between risk groups. Age, CD8^+^ Tregs, glial stem−like cells, and PD1^+^CD3^−^cells were consistently selected in sensitivity analyses and showed stable associations with OS (**Supplementary Figure 12**). More details regarding variable selection are provided in **Supplementary Note 2**.

In the grade 3 cohort, OS was positively associated with densities of CD68^−^ M2−like macrophages (HR = 0.37, 95% CI: 0.09−1.51, p = 0.167) and CD11b^−^CD14^+^ APCs (HR = 0.65, 95% CI: 0.23−1.89, p = 0.431), while age at diagnosis (HR = 1.3, 95% CI: 0.85−1.97, p = 0.224) was negatively associated with OS (**Supplementary Figure 11C**). Higher and lower risk groups in grade 3 glioma were separated by the fitted Cox regression model built using these multi−marker secondary phenotypes, with Kaplan−Meier analysis yielding a nonsignificant log−rank test (**Supplementary Figure 11C**, C−index = 0.671, 95% CI: 0.56−0.782; **Supplementary Figure 11D**, log−rank p = 0.192).

In the grade 2 cohort, all multi−marker secondary phenotypes present (non−zero) in at least 10% of samples were associated with hazard ratios greater than 1 in univariable Cox regression models. Neutrophils, microglia−like cells, MHC II^+^ neutrophils, MHC II^−^ monocytes, glial stem−like cells, CD68^−^ M2−like macrophages, CD163^+^ M2−like macrophages, CD206^+^ M2−like macrophages and CD11b^+^CD14^+^ APCs were association with poorer outcome (**Supplementary Figure 13**).

### Immune cell composition differences between glioma grades

We also examined differences in abundance of primary and multi−marker secondary phenotypes in grades 2-4 gliomas (methodological details in **Supplementary Note 3**). Among the 15 primary immune phenotypes, 8 exhibited grade−dependent increases. The primary phenotypes CD4^+^ and CD14^+^ showed significant differences across all pairwise comparisons among grade 2, grade 3, and grade 4 (**Supplementary Table 9**, **Supplementary Figure 14**). Expectedly, there were significant differences in age at diagnosis between glioma grades, progressively increasing from grade 2 to grade 3 to grade 4 (**Supplementary Figure 14**). The relative abundance of the CD20^+^ primary phenotype was significantly higher in grade 3 compared with grade 2 gliomas, while primary phenotypes CD15^+^ and CD163^+^ were more significantly more abundant in grade 4 relative to grade 3 glioma (**Supplementary Figure 14**). The CD3^+^, CD8^+^ and CD68^+^ primary phenotypes showed significant differences between grade 2 and grade 4 glioma (**Supplementary Figure 14**).

Several multi−marker secondary phenotypes were monotonically more abundant with increasing grade, including, innate−like T cells, double−negative T cells, cytotoxic T cells, T helper cells, double−positive T cells, Tregs, CD11b^−^CD14^+^ APCs, CD11b^+^CD14^+^ APCs, CD14^+^CD11b^−^MHC II^−^ cells, MHC II^−^ monocytes, glial stem−like cells, neutrophils, macrophages/microglia, M2 macrophages, and CD163^+^ M2−like macrophages (**Supplementary Figure 15**). Notably, T helper cells exhibited a significantly increasing trend, with significant differences observed across all pairwise comparisons (**Supplementary Figure 15**). Double−negative T cells, CD11b^−^CD14^+^ APCs, and CD11b^−^CD14^−^ APCs were significantly less abundant in grade 2 relative to grade 3 gliomas (**Supplementary Figure 15**). CD14^+^CD11b^−^MHC II^−^ cells, CD163^+^ M2−like macrophages, MHC II^−^ monocytes, and neutrophils were significantly less abundant in grade 3 compared with grade 4 (**Supplementary Figure 15**). Abundances of CD11b^+^CD14^+^ APCs, cytotoxic T cells, double−positive T cells, glial stem−like cells, innate−like T cells, M2 macrophages, macrophages/microglia, and Tregs were significantly different between grades 2 and 4 (**Supplementary Figure 15**). B cells were significantly less abundant in grade 2 relative to grades 3 and 4 (**Supplementary Figure 15**).

## Discussion

Molecular, genetic and protein tumor biomarkers to inform diagnosis, prognosis, and personalized therapy are typically based on tumor cell features.^1,29^ The concept of immune contexture was introduced so that the type, density, functional orientation and localization of immune cells within a tumor could inform patient management.^14^ We have characterized T, B, and myeloid cell infiltrates in tumor tissue taken at initial resection of newly−diagnosed glioma grades 2, 3 and 4 on the basis of the relative abundance (positive cells per 10,000 total cells) of specific immune phenotypes: (i) primary phenotypes defined as cells that were positive for the single immune markers (3×5 = 15 primary immune phenotypes from 3 mIF panels), (ii) secondary phenotypes defined on the basis of combinations of 5 immune−related markers in a given mIF panel (3×2^5^ = 96 secondary immune phenotypes from 3 mIF panels), and, (iii) 29 multi−marker phenotypes defined as biologically meaningful combinations of the 96 secondary immune phenotypes (**Table 2** and **Supplementary Table 2**). We identified four immune TME subtypes in grade 4 tumors (α, β, γ, δ) that were internally reproducible. Of note, grade 3 gliomas mapped onto these same four immune subtypes, with few outliers. Analogously, grade 2 gliomas could be mapped onto internally reproducible immune TME subtypes ε, ρ and τ that were defined on grade 3 gliomas, with no outliers. These findings suggest that immune TME subtypes α, β, γ and δ substantially capture the immune TME of all higher grade gliomas, and that immune TME subtypes ε, ρ and τ substantially capture the immune TME of both grade 2 and grade 3 gliomas.

G4α gliomas were characterized by high abundance of APCs and low levels of MHC II− monocytes, while G4β gliomas were high in regulatory T cells and myeloid cells but low in lymphocytes with effector functions. G4γ gliomas displayed high abundance of immune cell phenotypes, particularly lymphocytes with effector or helper functions, while G4δ gliomas were low in lymphoid and myeloid immune phenotypes and low in APCs. G4δ, G3ε, and G2φ tumors were characterized by low relative counts of lymphoid and myeloid immune phenotypes, with complete overlap between G2φ and G2ε immune subtypes, and almost complete overlap between G3ε and G3δ subtypes. These immune-poor subtypes tended to have the poorest outcomes. The immune subtype γ was characterized by high abundance of multiple immune phenotypes, particularly lymphocytes with effector or helper functions. These included NK cells, innate−like T cells, cytotoxic T cells, T helper cells, and double−positive T cells. In grade 3, G3γ had significantly longer OS than subtype G3δ, which was consistent with the OS in grade 4 subtype G4γ being longer than in subtype G4δ. Understanding the natural or therapy-driven progression, if any, of lower grade glioma immune subtypes into higher grade immune subtypes would require a longitudinal study.

Using unsupervised transcriptome analysis of TCGA grade 4 samples, Verhaak and colleagues defined four clusters, referred to as Classical, Mesenchymal, Neural, and Proneural, with Neural subtype being potentially non-tumor tissue, *IDH1* mutant disease being consistently classified as Proneural subtype, Classical subtype being most sensitive to therapy, and Mesenchymal subtype trending towards the poorest outcome.^30,31^ Glioma subtype was retained at recurrence in 55% of cases.^31^ Doucette et al. found that the mesenchymal subtype in TCGA grade 4 samples was enriched in genes reflective of antitumor proinflammatory adaptive and innate immune responses and immunosuppression.^32^ Kaffes et al. noted increased immune cell presence in Mesenchymal glioblastomas compared to Proneural and Classical subtypes.^33^ Martinez−Lage et al. found that macrophages and microglia were the predominant immune cell type in all grade 4 subtypes, with a significantly higher percentage in mesenchymal, followed by the classical and proneural subtypes.^34^ González−Tablas Pimenta et al. assayed single−cell suspensions from grade 4 samples by flow cytometry and found no significant differences in the relative distributions of immune cell subsets between grade 4 tissue specimens taken at diagnosis (*n*=44) and taken at recurrence (*n*=11).^35^

In our analysis, younger grade 4 patients experienced notably longer survival than older patients (**Supplementary Figure 11A**; **Supplementary Figure 12**), consistent with the known prognostic impact of age on patient survival.^36^ A significant proportion of immune cells were of myeloid origin in grade 4 gliomas, consistent with previous observations.^8^ The ratio of FoxP3^+^ to CD8^+^ cells has been described as a measure of the degree of immunosuppression.^13^ In our data, the ratio of FoxP3^+^ to CD8^+^ cells decreased with increasing glioma grade. PD−1^+^ cells were rare, in agreement with prior reports^11,37^. It has been suggested that low prevalence of PD−1^+^ T cells, particularly in the context of high prevalence of CD163^+^ cells, may reflect low activation of T cells in a highly immunosuppressive tumor microenvironment.^38^ Tregs have been reported to be rare in clinical gliomas^11,39^ and in preclinical glioma models^40^, with the evidence suggesting that M−2 macrophages and MDSCs are more important for the establishment of an immunosuppressive microenvironment in gliomas. In our grade 4 cohort, regulatory T cells (Tregs) were relatively rare, while macrophage/microglia/monocyte defined using markers from Panel 2 and Panel 3 were highly represented (**Supplementary Table 9**).

Klemm et al. noted that *IDH1−mut* gliomas were characterized by dominance of microglia with only low numbers of other immune cells, while *IDH1−wt* gliomas were characterized by an influx of monocyte−derived macrophages and neutrophils but low counts of lymphocytes.^39^ *IDH1−mut* status has also been reported to be associated with a quiescent immune profile.^37^ In contrast to several prior reports describing *IDH1*−associated differences in glioma immune infiltration, we did not observe statistically significant differences in marker densities between *IDH1−wt* and *IDH1−mut* tumors in our cohort (**Supplementary Table 10**). This difference may be attributable to our grade−stratified analytical approach, as the influence of *IDH1* status may be weaker within individual tumor grades. In addition, the limited number of *IDH1−mut* cases in the grade 4 cohort may have reduced the statistical power to detect *IDH1*−related differences. Consistent with reports from multiple groups studies,^11^ our data also demonstrated a tumor grade−specific increase in immune cell infiltration, with lower grade astrocytoma having sparse infiltrating immune cells and high grade astrocytoma characterized by abundant immune infiltrates (**Supplementary Figures 14 & 15**, **Supplementary Table 9**).

Li et al. classified low and high grade gliomas into four subtypes on the basis of expression levels of immune-related and metabolism-related genes, and found that grades 2 and 3 were defined by low metabolism and low infiltration of immune cells, while *IDH1-wt* grade 4 tumors were characterized by high metabolism and high immune infiltration.^41^ In a proteomic analysis of grades 2-4 gliomas, Zhang et al. identified two subtypes, a metabolic neural subgroup that is enriched in metabolic enzymes and neurotransmitter receptor proteins, and an immune subgroup that is marked by upregulation of immune and inflammatory proteins.^42^ Zeiner et al. surveyed glioma−associated microglia and macrophage (GAM) subpopulations in the microenvironments of low− and high−grade astrocytomas. Cells positive for the M2−associated marker CD206 were generally scarce in *IDH1−wt* grades 2−4, while CD163^+^ cells were somewhat more prevalent in *IDH1−wt* grade 4 and located mainly in perinecrotic areas with a non−significant increasing trend with increasing grade. Interestingly, in their analysis, high levels of CD68^+^ (>15%), CD163^+^ (>10%), and CD206^+^ (>3%) GAM subpopulations in the vital tumor core were associated with better survival in of *IDH1−wt* grade 4 patients.^43^ We performed a sensitivity analysis restricted to *IDH1−wt* grade 4 tumors in our data, which selected macrophage/microglia features in the multivariable Cox regression model, characterized by a CD68^+^CD163^−^CD206^−^ phenotype and this marker was associated with worse OS (**Supplementary Figure 12**).

There are limitations to prognostication based on analyses of immune infiltrates at single time points, such as in our current study. Our study also has several other limitations. The immune subtypes we have identified are internally reproducible, exploratory clusters, and validation on an independent external dataset will be required prior to any clinical interpretation of the identified immune TME subtypes. Our inferences were based on small 0.6 mm cores from original surgical tissue, though immune classifications were most often based on multiple cores (up to 3) per tumor to mitigate sampling error. The staining thresholds selected for each marker in each mIF panel were not 100% specific or sensitive for positivity/negativity, which effectively introduced some noise into the absolute and relative counts of specific phenotypes counted in each tumor core. Other significant limitations in our analysis include, zero−inflation in certain immune cell types, the lack of longitudinal or spatial analyses, and the fact that OS would be affected by the extent of surgical resection, the diversity of treatments received by individual patients, and molecular features such as MGMT promoter methylation status.

In summary, our analysis of mIF data identifies novel classifiers for glioma based on immune TME subtypes that predict patient survival independent of tumor grade and patient age. Immune cell density has been noted to be higher in the perivascular space of grade 4 glioma^12,38,39,43,46^, and has also been noted to be positively correlated with MRI−measured peritumoral edema in brain metastases^47^. We have previously identified an intratumoral “habitat” defined by hyperintensity on both contrast−enhanced T1−weighted MRI (indicative of the presence of tumor microvasculature) and Fluid−Attenuated Inversion Recovery MRI (indicative of the presence of edema) that is associated with long−term (≥ 36 months) survival post−diagnosis in newly−diagnosed glioblastoma.^45^ Our institution is prospectively collecting “geo−tagged” intraoperative tumor samples of grade 4 glioma and other brain tumors.^48^ In a planned study on this dataset we will investigate whether intratumoral habitats defined on pre−operative MRI can serve as surrogate markers of tumor immune TME subtypes. The immune TME in GBM is modulated by therapy and evolves during recurrence.^49^ An imaging biomarker of immune signatures would allow non−invasive, volumetric, longitudinal monitoring of changes and spatial heterogeneity in the glioma immune landscape, thereby supporting the development of novel therapies to guide favorable evolution of the immune microenvironment in high-grade glioma.

## Data Availability

All data produced in the present study are available upon reasonable request to the authors.

## Acknowledgments

The authors are grateful to all patients involved in the study, as well as their caregivers. They wish to acknowledge all of the treating clinicians, pathologists and research staff that contributed to the current study from the following centers: Moffitt Cancer Center in Tampa, FL; the Norton Cancer Institute in Louisville, KY; Morton Plant Hospital in Clearwater, FL; AdventHealth Cancer Institute in Orlando, FL; and Vanderbilt University Medical Center in Nashville, TN.

