## Supplementary Tables & Figures for "Immune Subtypes and Survival in Patients with Primary Glioma"

**Running title: Immune Subtypes in Primary Glioma**

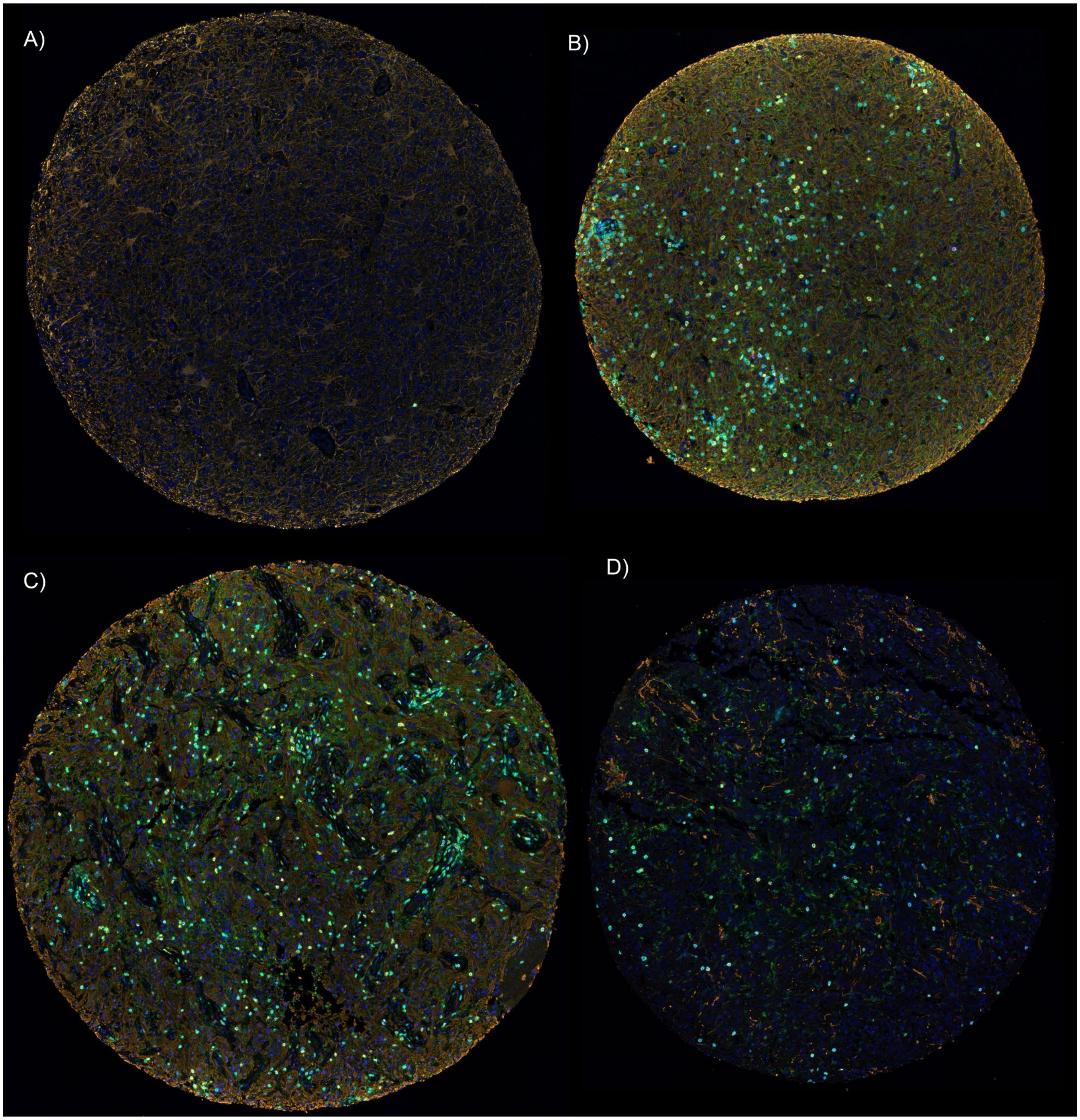


**Supplementary Figure 1.** Examples of ≈ 0.6 mm cores of glioma tissue stained with mIF Panel 1: FOXP3 (Red), CD3 (Cyan), CD4 (Green), CD8 (Yellow), PD-1 (Magenta), GFAP (Orange), and DAPI (Blue). **A)** Grade 4 example: all 5 markers are 0.0%; **B)** Grade 4 example: FOXP3 0.8%, CD3 11.2%, CD 2.6%, CD8 4.4%, PD1 0.3%; **C)** Grade 3 example: FOXP3 3.1%, CD3 12.1%, CD4 5.2%, CD8 4.8%, PD1 0.2%; **D)** Grade 1 example: FOXP3 0.1%, CD3 3.0%, CD4 10.6%, CD8 2.0%, PD1 0.0%.

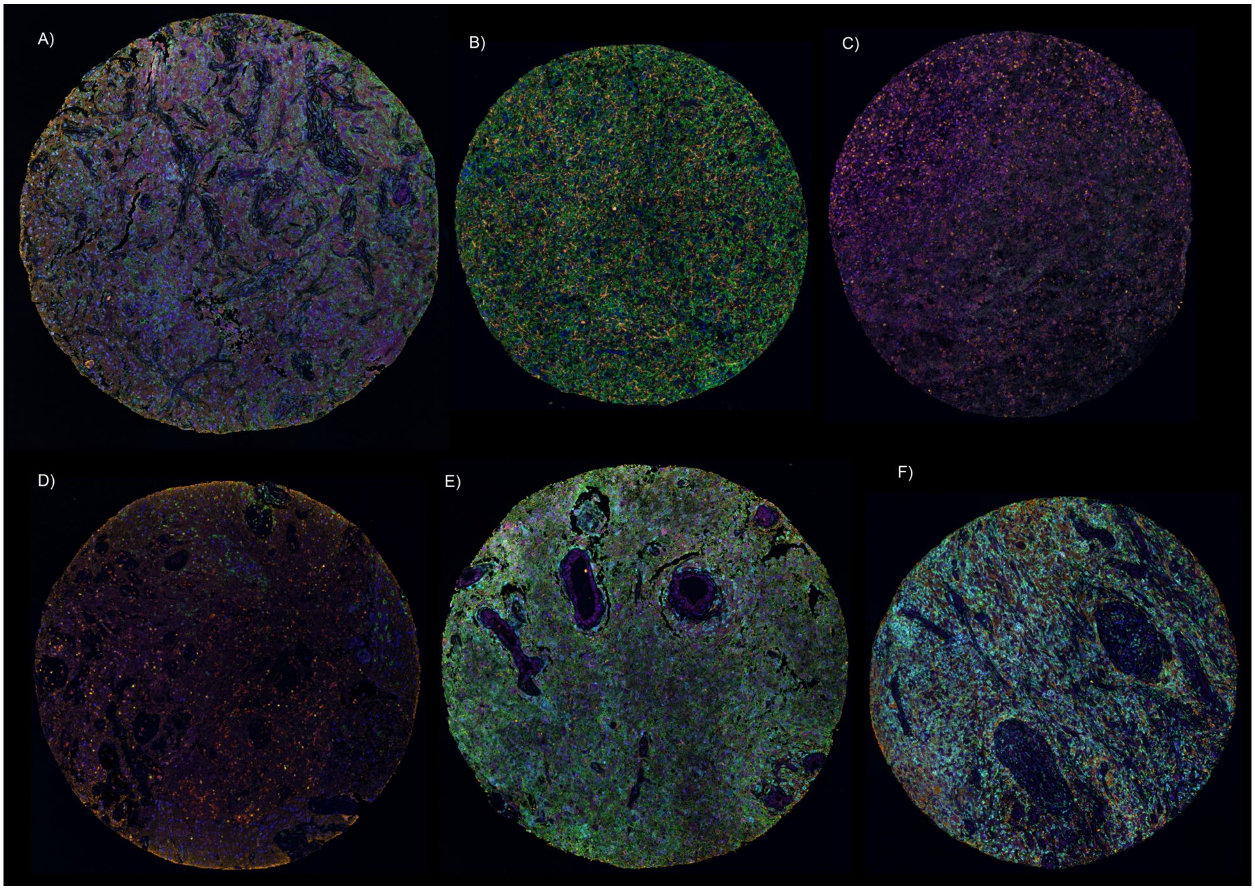


**Supplementary Figure 2.** Examples of ≈ 0.6 mm cores of glioma tissue stained with mIF Panel 2: CD11b (Red), MHCII (Cyan), CD14 (Green), CD15 (Yellow), CD33 (Magenta), GFAP (Orange), and DAPI (Blue). **A)** Grade 3 example: CD11b 14.1%, MHC II 20.6%, CD14 56.6%, CD15 0.1%, CD33 0.0%; **B)** Grade 4 example: CD14 70.7%, rest 0.0%; **C)** Grade 4 example: CD11b 58.3%, MHC II 0.1%, CD14 1.8%, CD15 29.4%, CD33 0.0%; **D)** Grade 4 example: CD11b 24.9%, MHC II 0.1%, CD14 9.7%, CD15 6.6%, CD33 0.0%; **E)** Grade 4 example: CD11b 0.7%, MHC II 74.2%, CD14 56.7%, CD15 0.0%, CD33 0.0%; **F)** Grade 4 example: CD11b 0.0%, MHC II 57.2%, CD14 51.1%, CD15 0.0%, CD33 0.0%.

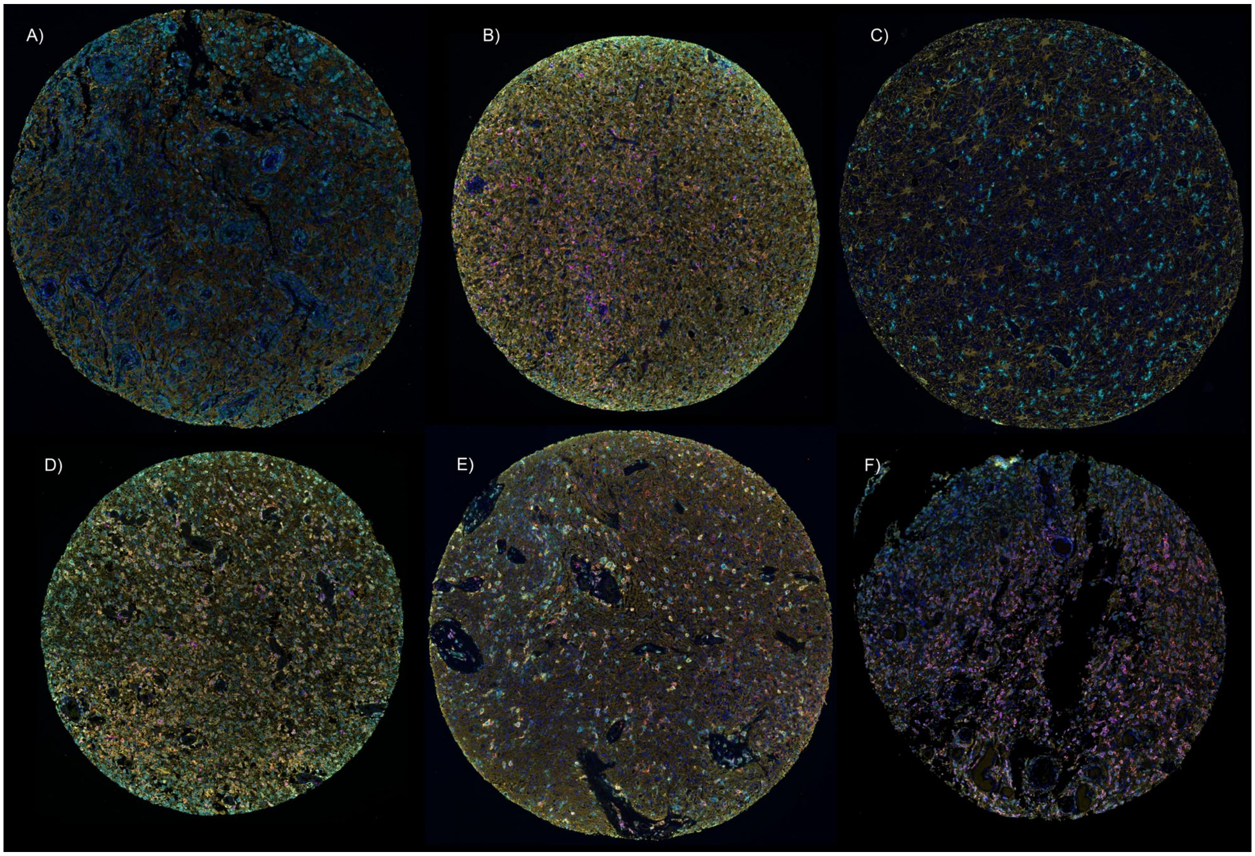


**Supplementary Figure 3.** Examples of ≈ 0.6 mm cores of glioma tissue stained with mIF Panel 3: CD163 (Red), CD68 (Cyan), CD20 (Green), CD19 (Yellow), CD206 (Magenta), GFAP (Orange), and DAPI (Blue). **A)** Grade 3 example: CD163 0.0%, CD68 60.2%, CD20 0.3%, CD19 0.0%, CD206 0.0%; **B)** Grade 4 example: CD163 11.9%, CD68 72.2%, CD20 0.1%, CD19 0.0%, CD206 3.7%; **C)** Grade 4 example: CD163 0.0%, CD68 11.0%, CD20 0.0%, CD19 0.0%, CD206 0.0%; **D)** Grade 4 example: CD163 30.2%, CD68 66.8%, CD20 0.2%, CD19 0.1%, CD206 4.1%; **E)** Grade 4 example: CD163 10.0%, CD68 39.0%, CD20 0.0%, CD19 0.0%, CD206 0.1%; **F)** Grade 4 example: CD163 7.9%, CD68 44.4%, CD20 0.1%, CD19 0.1%, CD206 10.7%.

| TMA | TMA1 | TMA1 | TMA2 | TMA2 | TMA3 | TMA4 | TMA4 | TMA4 | TMA5 | TMA6 |
|---|---|---|---|---|---|---|---|---|---|---|
| FOXP3 (OPAL 650) | −0.5 | −0.53 | −0.52 | −0.53 | 1.41 | −0.44 | −0.33 | 2.29 | −0.33 | −0.52 |
| CD3 (OPAL 690) | −0.05 | −0.57 | −0.49 | −0.64 | 2.71 | −0.46 | −0.27 | 0.24 | 0.08 | −0.56 |
| CD4 (OPAL 520) | −0.56 | −0.56 | −0.56 | −0.51 | 0.16 | −0.07 | 2.6 | 0.62 | −0.56 | −0.56 |
| CD8 (OPAL 570) | 2.06 | −0.85 | −0.34 | −0.97 | 0.21 | 0.39 | −0.14 | 1.2 | −0.97 | −0.6 |
| PD−1 (OPAL 540) | NA | NA | NA | NA | NA | NA | NA | NA | NA | NA |
| CD14 (OPAL 520) | 2.02 | −0.6 | 1.31 | −0.85 | −0.09 | −0.92 | −0.64 | 0.65 | −0.61 | −0.24 |
| CD15 (OPAL 570) | −0.43 | −0.43 | −0.43 | −0.43 | −0.43 | −0.43 | 2.61 | 0.83 | −0.43 | −0.43 |
| CD33 (OPAL 540) | −0.32 | −0.32 | −0.32 | −0.32 | −0.32 | −0.32 | 2.85 | −0.32 | −0.32 | −0.32 |
| CD11b (OPAL 650) | −0.4 | −0.39 | −0.37 | −0.4 | −0.16 | −0.29 | −0.15 | 2.83 | −0.29 | −0.39 |
| MHCII (OPAL 690) | −0.7 | −0.7 | −0.7 | −0.7 | 1.24 | −0.31 | 0.06 | 2.2 | 0.3 | −0.69 |
| CD19 (OPAL 570) | −0.32 | −0.32 | −0.32 | −0.32 | −0.32 | −0.32 | −0.32 | −0.32 | 2.85 | −0.32 |
| CD20 (OPAL 520) | −0.32 | −0.32 | −0.32 | 2.85 | −0.32 | −0.32 | −0.32 | −0.32 | −0.32 | −0.32 |
| CD206 (OPAL 540) | −0.42 | −0.42 | −0.4 | −0.23 | −0.47 | −0.42 | −0.47 | −0.19 | 0.25 | 2.78 |
| CD163 (OPAL 620) | 0.1 | −0.34 | −0.37 | −0.38 | −0.31 | −0.38 | −0.38 | −0.38 | 2.81 | −0.38 |
| CD68 (OPAL 690) | 1.8 | −0.61 | 0.31 | −0.76 | −0.44 | −1.15 | −0.82 | −0.32 | 0.54 | 1.46 |

**Supplementary Table 1:** Z−scores of marker expression in Tonsil control tissue across 6 TMAs.

We assessed whether any TMA behaved as an outlier by comparing the proportion of positive cells across all markers.

**Panel 1**

| | NK Cells | innate−like T cells | double−negative T cells | cytotoxic T cells | T helper cells | double−positive T cells | PD1$^{+}$CD3$^{-}$ cells | PD1$^{+}$ T lymphocytes | FoxP3$^{+}$CD3$^{-}$ cells | double−negative Tregs | CD8$^{+}$ Tregs | Tregs |
|---|---|---|---|---|---|---|---|---|---|---|---|---|
| $CD3^{-}CD4^{-}CD8^{+}FOXP3^{-}PD1^{-}$ | + | | | | | | | | | | | |
| $CD3^{-}CD4^{+}CD8^{-}FOXP3^{-}PD1^{-}$ | | + | | | | | | | | | | |
| $CD3^{-}CD4^{+}CD8^{+}FOXP3^{-}PD1^{-}$ | | + | | | | | | | | | | |
| $CD3^{+}CD4^{-}CD8^{-}FOXP3^{-}PD1^{-}$ | | | + | | | | | | | | | |
| $CD3^{+}CD4^{-}CD8^{+}FOXP3^{-}PD1^{-}$ | | | | + | | | | | | | | |
| $CD3^{+}CD4^{+}CD8^{-}FOXP3^{-}PD1^{-}$ | | | | | + | | | | | | | |
| $CD3^{+}CD4^{+}CD8^{+}FOXP3^{-}PD1^{-}$ | | | | | | + | | | | | | |
| $CD3^{-}CD4^{-}CD8^{-}FOXP3^{-}PD1^{+}$ | | | | | | | + | | | | | |
| $CD3^{-}CD4^{-}CD8^{+}FOXP3^{-}PD1^{+}$ | | | | | | | + | | | | | |
| $CD3^{-}CD4^{+}CD8^{-}FOXP3^{-}PD1^{+}$ | | | | | | | + | | | | | |
| $CD3^{+}CD4^{-}CD8^{-}FOXP3^{-}PD1^{+}$ | | | | | | | | + | | | | |
| $CD3^{+}CD4^{-}CD8^{+}FOXP3^{-}PD1^{+}$ | | | | | | | | + | | | | |
| $CD3^{+}CD4^{+}CD8^{+}FOXP3^{-}PD1^{+}$ | | | | | | | | + | | | | |
| $CD3^{+}CD4^{-}CD8^{-}FOXP3^{+}PD1^{+}$ | | | | | | | | + | | | | |
| $CD3^{-}CD4^{-}CD8^{-}FOXP3^{+}PD1^{-}$ | | | | | | | | | + | | | |
| $CD3^{-}CD4^{-}CD8^{+}FOXP3^{+}PD1^{-}$ | | | | | | | | | + | | | |
| $CD3^{-}CD4^{+}CD8^{-}FOXP3^{+}PD1^{-}$ | | | | | | | | | + | | | |
| $CD3^{+}CD4^{-}CD8^{-}FOXP3^{+}PD1^{-}$ | | | | | | | | | | + | | |
| $CD3^{+}CD4^{-}CD8^{+}FOXP3^{+}PD1^{-}$ | | | | | | | | | | | + | |
| $CD3^{+}CD4^{+}CD8^{-}FOXP3^{+}PD1^{-}$ | | | | | | | | | | | | + |
| $CD3^{+}CD4^{+}CD8^{+}FOXP3^{+}PD1^{-}$ | | | | | | | | | | | | + |

**Panel 2**

| | CD11b$^{-}$CD14$^{-}$ APCs | CD11b$^{-}$CD14$^{+}$ APCs | CD11b$^{+}$CD14$^{+}$ APCs | activated microglia | MHC II$^{+}$ neutrophils | CD14$^{+}$CD11b$^{-}$ MHC II$^{-}$ cells | MHC II$^{-}$ monocytes | myeloid-like cells | glial stem−like cells | microglia−like cells | neutrophils |
|---|---|---|---|---|---|---|---|---|---|---|---|
| $CD11b^{-}CD14^{-}CD15^{-}CD33^{-}MHCII^{+}$ | + | | | | | | | | | | |
| $CD11b^{-}CD14^{-}CD15^{-}CD33^{+}MHCII^{+}$ | + | | | | | | | | | | |
| $CD11b^{-}CD14^{-}CD15^{+}CD33^{-}MHCII^{+}$ | + | | | | | | | | | | |
| $CD11b^{-}CD14^{-}CD15^{+}CD33^{+}MHCII^{+}$ | + | | | | | | | | | | |
| $CD11b^{-}CD14^{+}CD15^{-}CD33^{-}MHCII^{+}$ | | + | | | | | | | | | |
| $CD11b^{-}CD14^{+}CD15^{-}CD33^{+}MHCII^{+}$ | | + | | | | | | | | | |
| $CD11b^{-}CD14^{+}CD15^{+}CD33^{-}MHCII^{+}$ | | + | | | | | | | | | |
| $CD11b^{+}CD14^{+}CD15^{-}CD33^{-}MHCII^{+}$ | | | + | | | | | | | | |
| $CD11b^{+}CD14^{+}CD15^{-}CD33^{+}MHCII^{+}$ | | | + | | | | | | | | |
| $CD11b^{+}CD14^{+}CD15^{+}CD33^{-}MHCII^{+}$ | | | + | | | | | | | | |
| $CD11b^{+}CD14^{+}CD15^{+}CD33^{+}MHCII^{+}$ | | | + | | | | | | | | |
| $CD11b^{+}CD14^{-}CD15^{-}CD33^{-}MHCII^{+}$ | | | | + | | | | | | | |
| $CD11b^{+}CD14^{-}CD15^{-}CD33^{+}MHCII^{+}$ | | | | + | | | | | | | |
| $CD11b^{+}CD14^{-}CD15^{+}CD33^{-}MHCII^{+}$ | | | | | + | | | | | | |
| $CD11b^{+}CD14^{-}CD15^{+}CD33^{+}MHCII^{+}$ | | | | | + | | | | | | |
| $CD11b^{-}CD14^{+}CD15^{-}CD33^{-}MHCII^{-}$ | | | | | | + | | | | | |
| $CD11b^{-}CD14^{+}CD15^{-}CD33^{+}MHCII^{-}$ | | | | | | + | | | | | |
| $CD11b^{-}CD14^{+}CD15^{+}CD33^{-}MHCII^{-}$ | | | | | | + | | | | | |
| $CD11b^{+}CD14^{+}CD15^{-}CD33^{-}MHCII^{-}$ | | | | | | | + | | | | |
| $CD11b^{+}CD14^{+}CD15^{-}CD33^{+}MHCII^{-}$ | | | | | | | + | | | | |
| $CD11b^{+}CD14^{+}CD15^{+}CD33^{-}MHCII^{-}$ | | | | | | | + | | | | |
| $CD11b^{-}CD14^{-}CD15^{-}CD33^{+}MHCII^{-}$ | | | | | | | | + | | | |
| $CD11b^{-}CD14^{-}CD15^{+}CD33^{+}MHCII^{-}$ | | | | | | | | + | | | |
| $CD11b^{+}CD14^{-}CD15^{-}CD33^{+}MHCII^{-}$ | | | | | | | | + | | | |
| $CD11b^{-}CD14^{-}CD15^{+}CD33^{-}MHCII^{-}$ | | | | | | | | | + | | |
| $CD11b^{+}CD14^{-}CD15^{-}CD33^{-}MHCII^{-}$ | | | | | | | | | | + | |
| $CD11b^{+}CD14^{-}CD15^{+}CD33^{-}MHCII^{-}$ | | | | | | | | | | | + |

**Panel 3**

| | B cells | macrophages/microglia | M2 macrophages | CD206$^{+}$ M2−like macrophages | CD163$^{+}$ M2−like macrophages | CD68$^{-}$ M2−like macrophages |
|---|---|---|---|---|---|---|
| $CD68^{-}CD163^{-}CD206^{-}CD19^{+}CD20^{-}$ | + | | | | | |
| $CD68^{-}CD163^{-}CD206^{+}CD19^{+}CD20^{-}$ | + | | | | | |
| $CD68^{-}CD163^{+}CD206^{-}CD19^{+}CD20^{-}$ | + | | | | | |
| $CD68^{-}CD163^{+}CD206^{+}CD19^{+}CD20^{-}$ | + | | | | | |
| $CD68^{-}CD163^{-}CD206^{-}CD19^{-}CD20^{+}$ | + | | | | | |

| | | | | | | |
|---|---|---|---|---|---|---|
| CD68$^{-}$CD163$^{+}$CD206$^{-}$CD19$^{-}$CD20$^{+}$ | + | | | | | |
| CD68$^{-}$CD163$^{-}$CD206$^{+}$CD19$^{-}$CD20$^{+}$ | + | | | | | |
| CD68$^{+}$CD163$^{-}$CD206$^{-}$CD19$^{+}$CD20$^{-}$ | | + | | | | |
| CD68$^{+}$CD163$^{-}$CD206$^{-}$CD19$^{-}$CD20$^{-}$ | | + | | | | |
| CD68$^{+}$CD163$^{-}$CD206$^{-}$CD19$^{-}$CD20$^{+}$ | | + | | | | |
| CD68$^{+}$CD163$^{+}$CD206$^{+}$CD19$^{-}$CD20$^{-}$ | | | + | | | |
| CD68$^{+}$CD163$^{+}$CD206$^{+}$CD19$^{+}$CD20$^{-}$ | | | + | | | |
| CD68$^{+}$CD163$^{+}$CD206$^{+}$CD19$^{-}$CD20$^{+}$ | | | + | | | |
| CD68$^{+}$CD163$^{-}$CD206$^{+}$CD19$^{-}$CD20$^{-}$ | | | | + | | |
| CD68$^{+}$CD163$^{-}$CD206$^{+}$CD19$^{+}$CD20$^{-}$ | | | | + | | |
| CD68$^{+}$CD163$^{-}$CD206$^{+}$CD19$^{-}$CD20$^{+}$ | | | | + | | |
| CD68$^{+}$CD163$^{+}$CD206$^{-}$CD19$^{-}$CD20$^{-}$ | | | | | + | |
| CD68$^{+}$CD163$^{+}$CD206$^{-}$CD19$^{+}$CD20$^{-}$ | | | | | + | |
| CD68$^{-}$CD163$^{-}$CD206$^{+}$CD19$^{-}$CD20$^{-}$ | | | | | | + |
| CD68$^{-}$CD163$^{+}$CD206$^{+}$CD19$^{-}$CD20$^{-}$ | | | | | | + |
| CD68$^{-}$CD163$^{+}$CD206$^{-}$CD19$^{-}$CD20$^{-}$ | | | | | | + |

**Supplementary Table 2:** Multi−marker secondary phenotypes.

| Grade 4 cohort | | | | | | | | |
|---|---|---|---|---|---|---|---|---|
| Principal component | Standard deviation | Cumulative proportion | Principal component | Standard deviation | Cumulative proportion | Principal component | Standard deviation | Cumulative proportion |
| *PC1* | 3.14 | 0.14 | *PC9* | 1.58 | 0.56 | *PC17* | 1.75 | 0.78 |
| *PC2* | 2.27 | 0.22 | *PC10* | 1.53 | 0.60 | *PC18* | 1.14 | 0.80 |
| *PC3* | 2.16 | 0.29 | *PC11* | 1.47 | 0.63 | *PC19* | 1.09 | 0.82 |
| *PC4* | 2.02 | 0.35 | *PC12* | 1.43 | 0.66 | *PC20* | 1.04 | 0.83 |
| *PC5* | 1.89 | 0.40 | *PC13* | 1.42 | 0.69 | *PC21* | 1.02 | 0.85 |
| *PC6* | 1.82 | 0.44 | *PC14* | 1.39 | 0.71 | *PC22* | 1.01 | 0.86 |
| *PC7* | 1.72 | 0.49 | *PC15* | 1.29 | 0.73 | | | |
| *PC8* | 1.61 | 0.53 | *PC16* | 1.25 | 0.76 | | | |
| **Grade 3 cohort** | | | | | | | | |
| Principal component | Standard deviation | Cumulative proportion | Principal component | Standard deviation | Cumulative proportion | Principal component | Standard deviation | Cumulative proportion |
| *PC1* | 3.31 | 0.20 | *PC6* | 1.79 | 0.63 | *PC11* | 1.23 | 0.80 |
| *PC2* | 2.57 | 0.32 | *PC7* | 1.56 | 0.67 | *PC12* | 1.14 | 0.83 |
| *PC3* | 2.32 | 0.42 | *PC8* | 1.46 | 0.71 | *PC13* | 1.12 | 0.85 |
| *PC4* | 2.13 | 0.50 | *PC9* | 1.39 | 0.75 | *PC14* | 1.06 | 0.87 |
| *PC5* | 1.97 | 0.57 | *PC10* | 1.26 | 0.77 | *PC15* | 1.02 | 0.89 |
| **Grade 2 cohort** | | | | | | | | |
| Principal component | Standard deviation | Cumulative proportion | Principal component | Standard deviation | Cumulative proportion | Principal component | Standard deviation | Cumulative proportion |
| *PC1* | 3.99 | 0.32 | *PC5* | 1.71 | 0.76 | *PC9* | 1.05 | 0.89 |
| *PC2* | 3.02 | 0.51 | *PC6* | 1.51 | 0.82 | *PC10* | 1.03 | 0.91 |
| *PC3* | 2.25 | 0.61 | *PC7* | 1.29 | 0.84 | *PC11* | 1.00 | 0.93 |
| *PC4* | 2.01 | 0.70 | *PC8* | 1.22 | 0.87 | | | |

**Supplementary Table 3:** Proportion of variance explained by each selected principal component (Kaiser criterion, eigenvalue > 1) within each tumor grade cohort. For each PC, the table reports the mean proportion of explained variance across samples and the corresponding standard deviation.

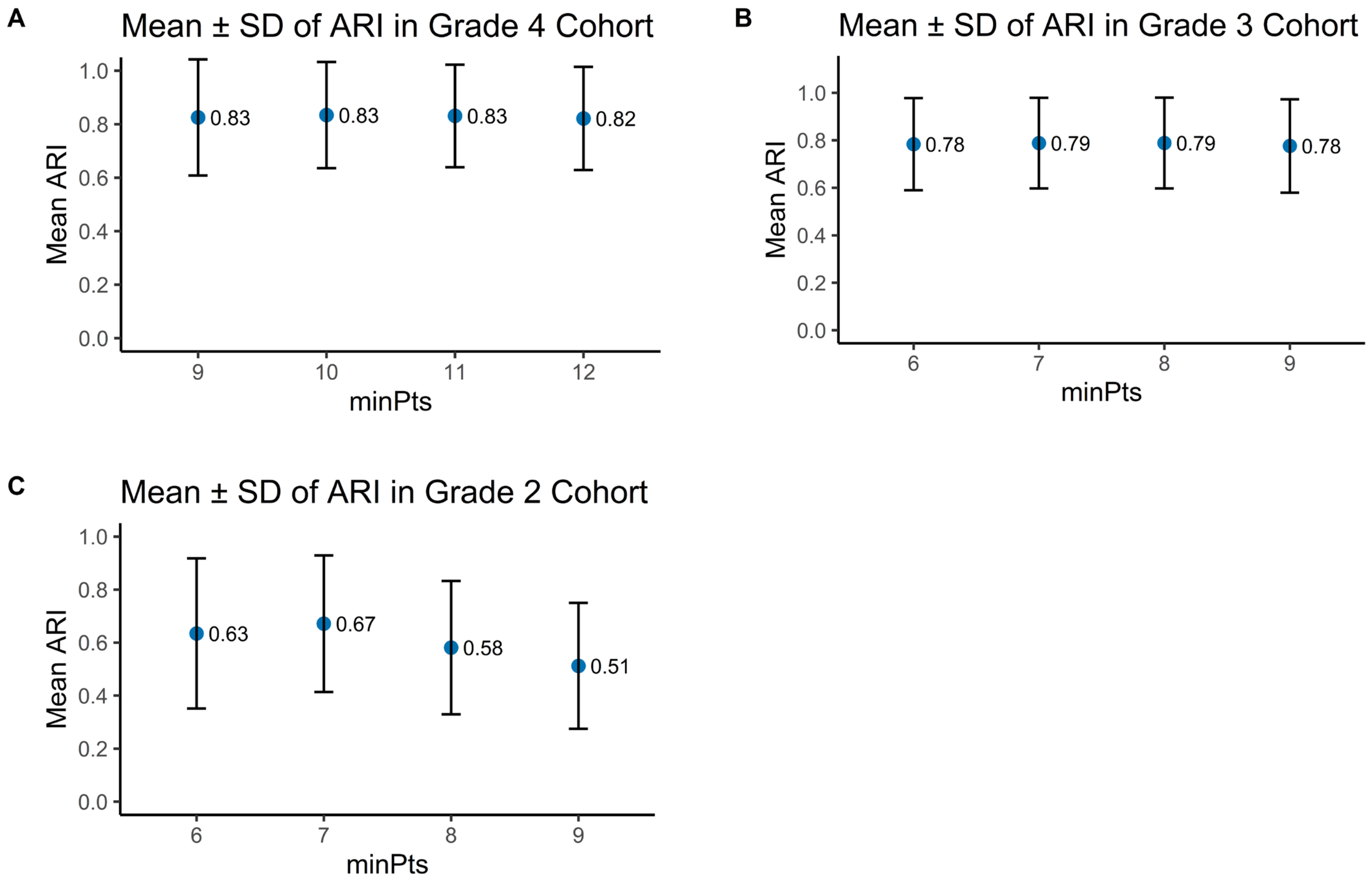


**Supplementary Figure 4:** Stability analysis of clustering results across four different minPts settings. **(A)** Grade 4 cohort (corresponding to Main Figure 1A), **(B)** Grade 3 cohort (corresponding to Main Figure 1C), and **(C)** Grade 2 cohort (corresponding to Main Figure 1E). For each minPts value, clustering was repeated using 3,001 random seeds (0-3000). Bars indicate the mean Adjusted Rand Index (ARI), with error bars representing the standard deviation, summarizing clustering consistency across runs.

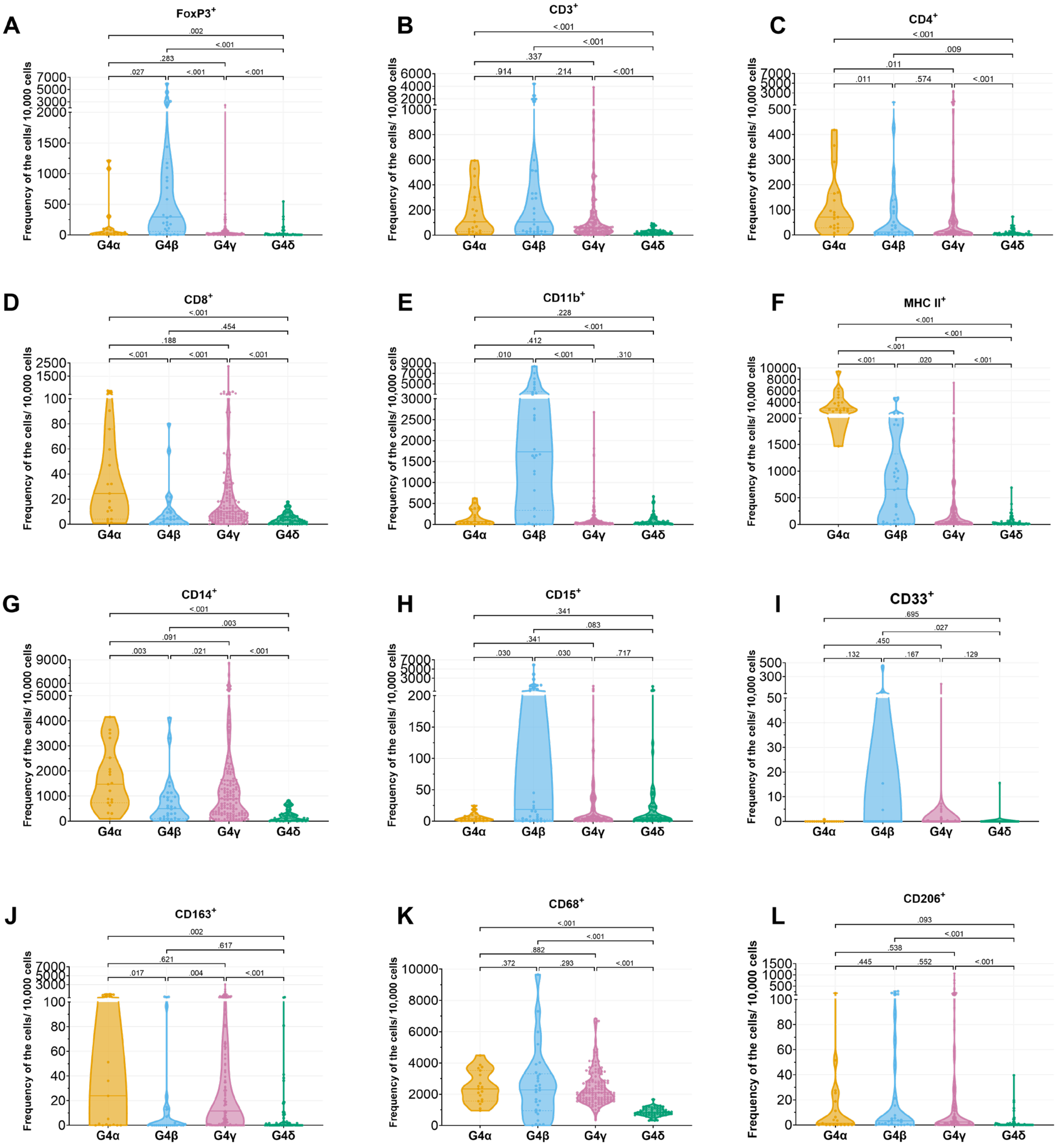


**Supplementary Figure 5:** Violin plots of primary phenotypes significant by Kruskal−Wallis testing in grade 4 cohort, stratified by subtypes.

| Cell types | Mean (SD) | | | | Median (IQR) | | | | Adjusted *P* values |
|---|---|---|---|---|---|---|---|---|---|
| | G4α (*n* = 19) | G4β (*n* = 30) | G4γ (*n* = 135) | G4δ (*n* = 56) | G4α (*n* = 19) | G4β (*n* = 30) | G4γ (*n* = 135) | G4δ (*n* = 56) | |
| All T cells (CD3$^+$, regardless of other markers in Panel 1) | 1.85 (1.89) | 5.44 (9.9) | 1.52 (3.68) | 0.24 (0.22) | 1.06 (0.32−2.91) | 1.05 (0.29−4.67) | 0.61 (0.29−1.28) | 0.16 (0.1−0.31) | **< 0.001** |
| Regulatory T cells (Tregs, CD3$^+$FOXP3$^+$, regardless of other markers in Panel 1) | 0.31 (0.55) | 3.18 (6.02) | 0.16 (0.43) | 0.04 (0.1) | 0.14 (0.03−0.27) | 0.19 (0.03−1.86) | 0.04 (0.01−0.13) | 0 (0−0.04) | **< 0.001** |
| T helper cells (CD3$^+$CD4$^+$, regardless of other markers in Panel 1) | 0.36 (0.37) | 0.22 (0.41) | 0.37 (0.77) | 0.04 (0.05) | 0.22 (0.09−0.6) | 0.05 (0.01−0.15) | 0.07 (0.02−0.31) | 0.02 (0−0.05) | **< 0.001** |
| Cytotoxic T cells (CD3$^+$CD8$^+$, regardless of other markers in Panel 1) | 0.6 (0.93) | 0.09 (0.16) | 0.4 (1.9) | 0.04 (0.04) | 0.23 (0.06−0.61) | 0.03 (0−0.09) | 0.09 (0.04−0.25) | 0.03 (0.01−0.05) | **< 0.001** |
| Antigen−Presenting cells (APCs, MHCII$^+$CD14$^-$CD11b$^-$ or MHCII$^+$CD14$^+$CD11b$^-$ or MHCII$^+$CD14$^+$CD11b$^+$ or MHCII$^+$CD14$^-$CD11b$^+$, regardless of other markers in Panel 2) | 37.87 (19.01) | 9.46 (12.32) | 2.99 (7.46) | 0.58 (1.15) | 29.85 (26.18−43.97) | 6.61 (0.3−11.2) | 0.55 (0.13−2.84) | 0.12 (0.05−0.57) | **< 0.001** |
| Macrophages/microglia/monocytes (CD14$^+$CD11b$^-$ or CD14$^+$CD11b$^+$ or CD15$^+$ or CD11b$^+$, regardless of other markers in Panel 2) | 18.43 (13.26) | 38.33 (37.5) | 15.23 (17.82) | 3.55 (3.64) | 15.72 (8.39−24.01) | 27.95 (12.89−51.18) | 9.98 (3.88−18.3) | 2.35 (0.69−5.03) | **< 0.001** |
| Non−M2 Macrophages/microglia/monocytes (CD68$^+$CD163$^-$CD206$^-$ or CD68$^-$CD163$^-$CD206$^+$ or CD68$^-$CD163$^+$CD206$^-$, regardless of other markers in Panel 3) | 23.15 (9.04) | 25.01 (21.86) | 23.19 (9.74) | 8.45 (2.68) | 23.18 (15.59−28.82) | 22.7 (9.73−30.56) | 20.78 (16.65−28.22) | 8.23 (6.88−10.28) | **< 0.001** |
| All M2− macrophages (CD68$^+$CD163$^+$CD206$^+$ or CD68$^+$CD163$^-$CD206$^+$ or CD68$^+$CD163$^+$CD206$^-$, regardless of other markers in Panel 3) | 1.86 (2.79) | 0.5 (0.92) | 1.61 (3.98) | 0.11 (0.29) | 0.24 (0.01−2.73) | 0.03 (0−0.53) | 0.21 (0.02−1.3) | 0 (0−0.08) | **< 0.001** |

**Supplementary Table 4:** Major immune cell phenotypes in grade 4 by tumor subtypes.

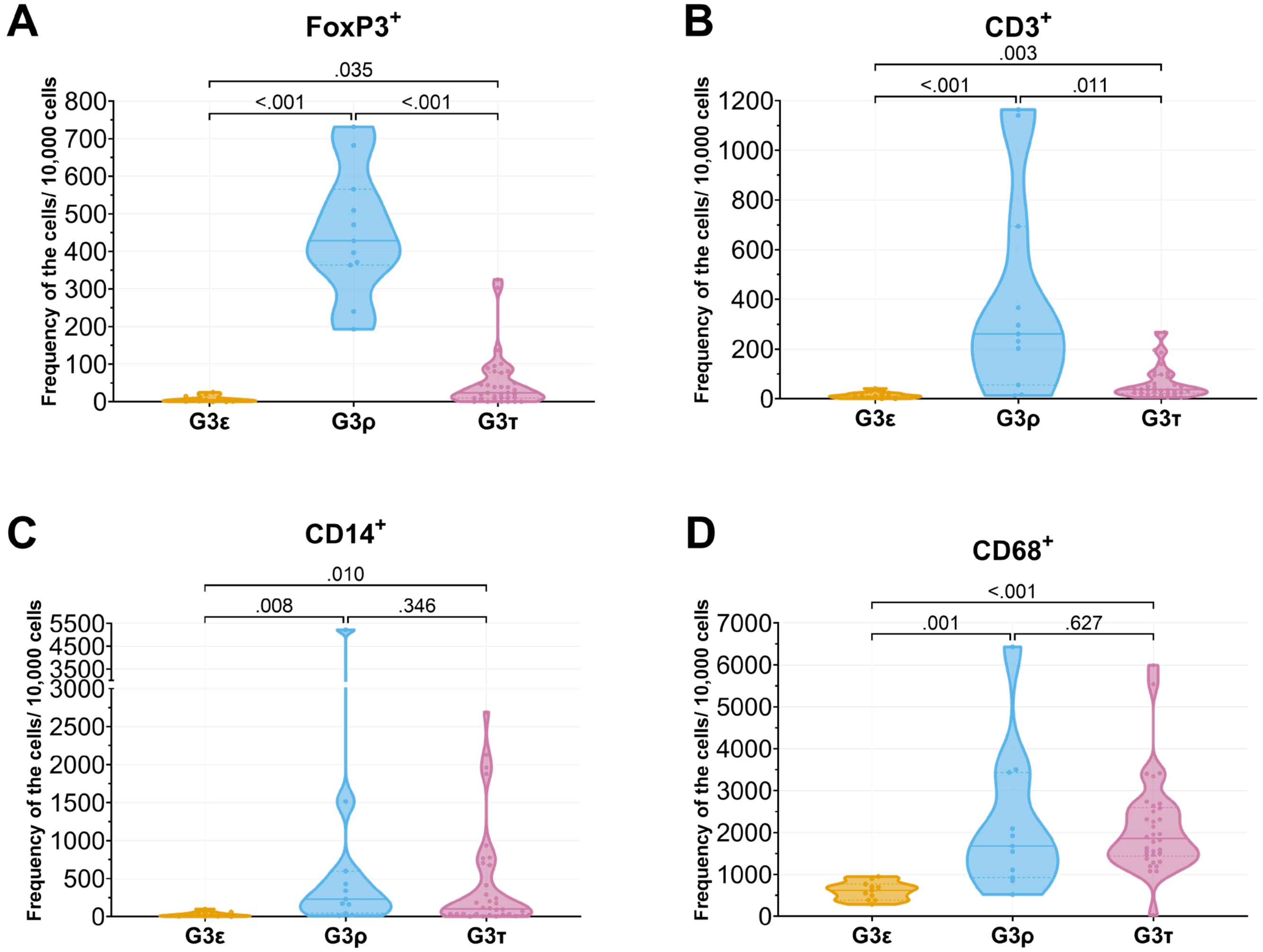


**Supplementary Figure 6:** Violin plots of primary phenotypes significant by Kruskal−Wallis testing in grade 3 cohort, stratified by subtypes.

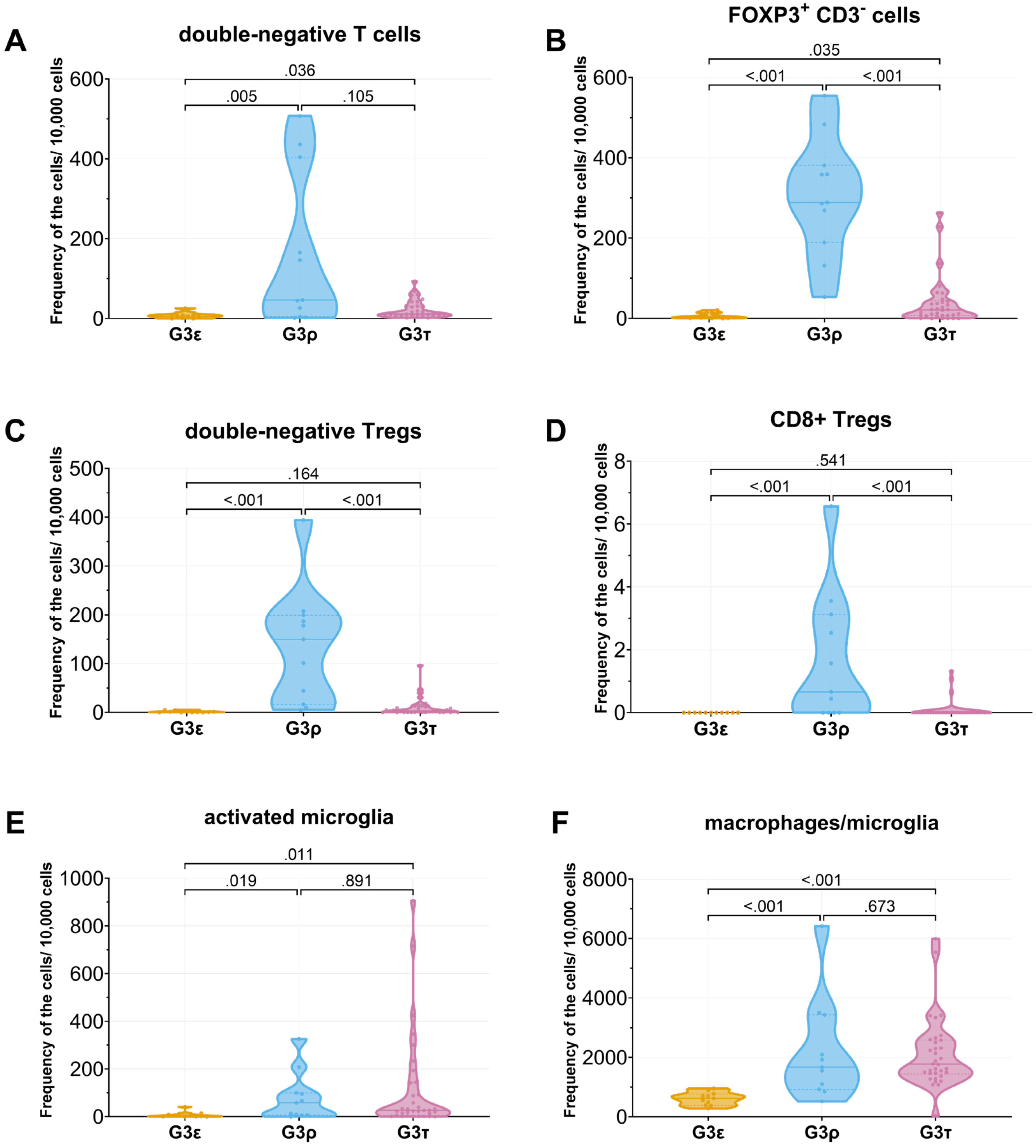


**Supplementary Figure 7:** Violin plots of multi-marker secondary phenotypes significant by Kruskal−Wallis testing in grade 3 cohort, stratified by subtypes. Dunn's post hoc tests were applied, with FDR−adjusted p−values reported.

| Cell types | Mean (SD) | | | Median (IQR) | | | Adjusted *P* values |
|---|---|---|---|---|---|---|---|
| | G3ε (*n* = 11) | G3ρ (*n* = 11) | G3τ (*n* = 34) | G3ε (*n* = 11) | G3ρ (*n* = 11) | G3τ (*n* = 34) | |
| All T cells ($CD3^+$, regardless of other markers in Panel 1) | 0.13 (0.12) | 4.04 (4.16) | 0.66 (0.7) | 0.11 (0.05−0.18) | 2.61 (1.29–5.31) | 0.38 (0.23−0.93) | **<0.001** |
| Regulatory T cells (Tregs, $CD3^+FOXP3^+$, regardless of other markers in Panel 1) | 0.01 (0.02) | 1.45 (1.25) | 0.12 (0.2) | 0.01 (0–0.02) | 1.5 (0.54–1.93) | 0.05 (0−0.14) | **<0.001** |
| T helper cells ($CD3^+CD4^+$, regardless of other markers in Panel 1) | 0.02 (0.04) | 0.66 (1.36) | 0.2 (0.47) | 0 (0−0.01) | 0.04 (0–0.17) | 0.02 (0−0.08) | 0.159 |
| Cytotoxic T cells ($CD3^+CD8^+$, regardless of other markers in Panel 1) | 0.02 (0.04) | 0.41 (1.08) | 0.16 (0.27) | 0 (0−0.02) | 0.05 (0.03−0.18) | 0.07 (0.01−0.22) | **0.017** |
| Antigen−Presenting cells (APCs, $MHCII^+CD14^-CD11b^-$ or $MHCII^+CD14^+CD11b^-$ or $MHCII^+CD14^+CD11b^+$ or $MHCII^+CD14^-CD11b^+$, regardless of other markers in Panel 2) | 0.52 (0.72) | 7.03 (9.33) | 7.11 (12.44) | 0.2 (0.1−0.65) | 1.17 (0.32−12.97) | 1.45 (0.21−6.73) | **0.043** |
| Macrophages/microglia/monocytes ($CD14^+CD11b^-$ or $CD14^+CD11b^+$ or $CD15^+$ or $CD11b^+$, regardless of other markers in Panel 2) | 0.66 (0.29) | 11.36 (18.81) | 8.95 (11.74) | 0.6 (0.45−0.85) | 4.15 (1.66−10.44) | 3.62 (0.92−13.74) | **0.004** |
| Non−M2 Macrophages/microglia/monocytes ($CD68^+CD163^-CD206^-$ or $CD68^-CD163^-CD206^+$ or $CD68^-CD163^+CD206^-$, regardless of other markers in Panel 3) | 6.1 (2.16) | 21.8 (17.1) | 21.19 (11.81) | 6.19 (4.42−7.31) | 16.69 (10.15−27.61) | 17.94 (14.66−25.63) | **<0.001** |
| All M2− macrophages ($CD68^+CD163^+CD206^+$ or $CD68^+CD163^-CD206^+$ or $CD68^+CD163^+CD206^-$, regardless of other markers in Panel 3) | 0.03 (0.04) | 0.04 (0.04) | 0.21 (0.6) | 0 (0−0.06) | 0.01 (0−0.07) | 0.01 (0−0.2) | 0.611 |

**Supplementary Table 5:** Major immune cell phenotypes in grade 3 by tumor subtypes.

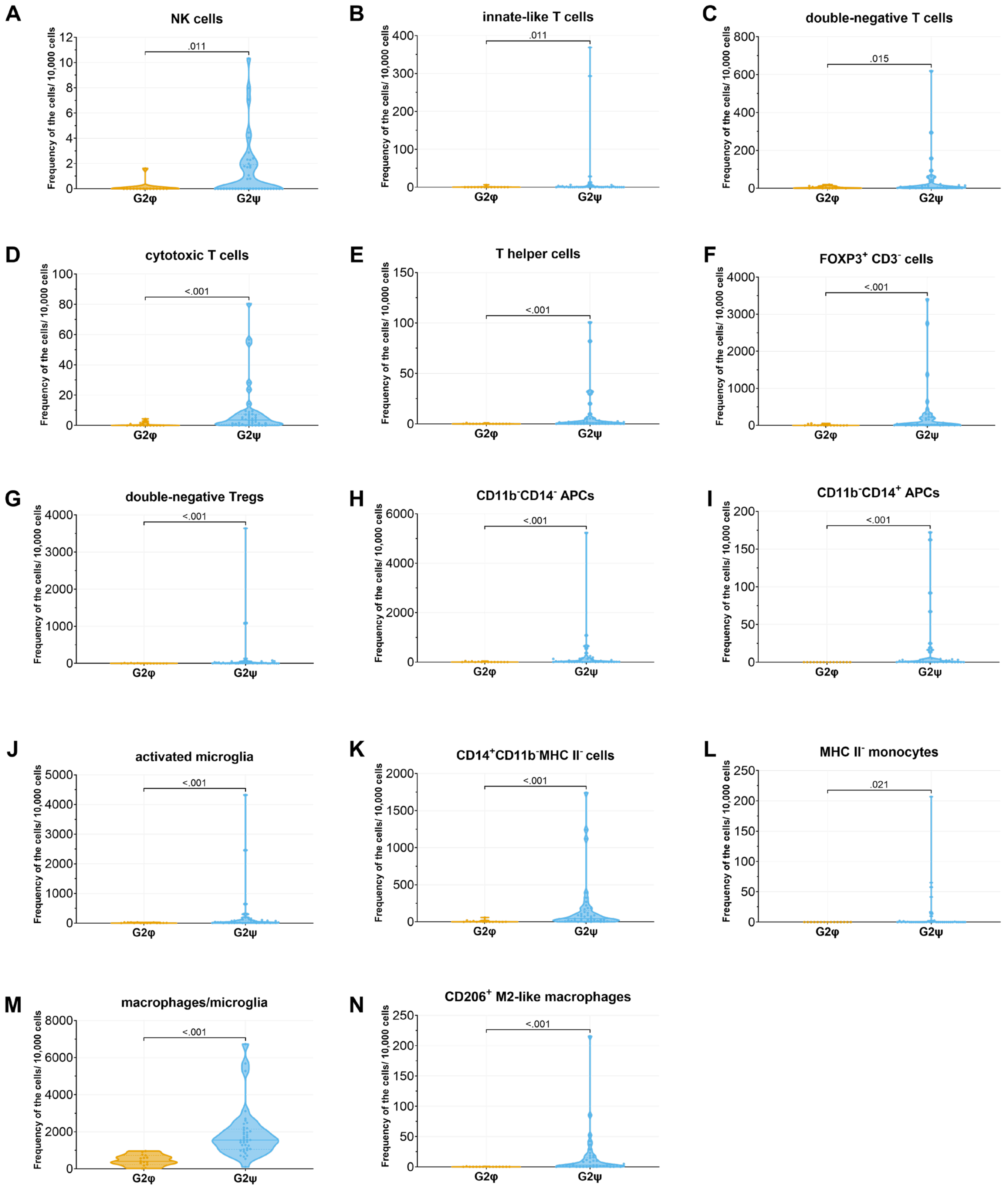


**Supplementary Figure 8:** Violin plots of multi-marker secondary phenotypes significant by Kruskal−Wallis testing in grade 2 cohort, stratified by subtypes. Dunn's post hoc tests were applied, with FDR−adjusted p−values reported.

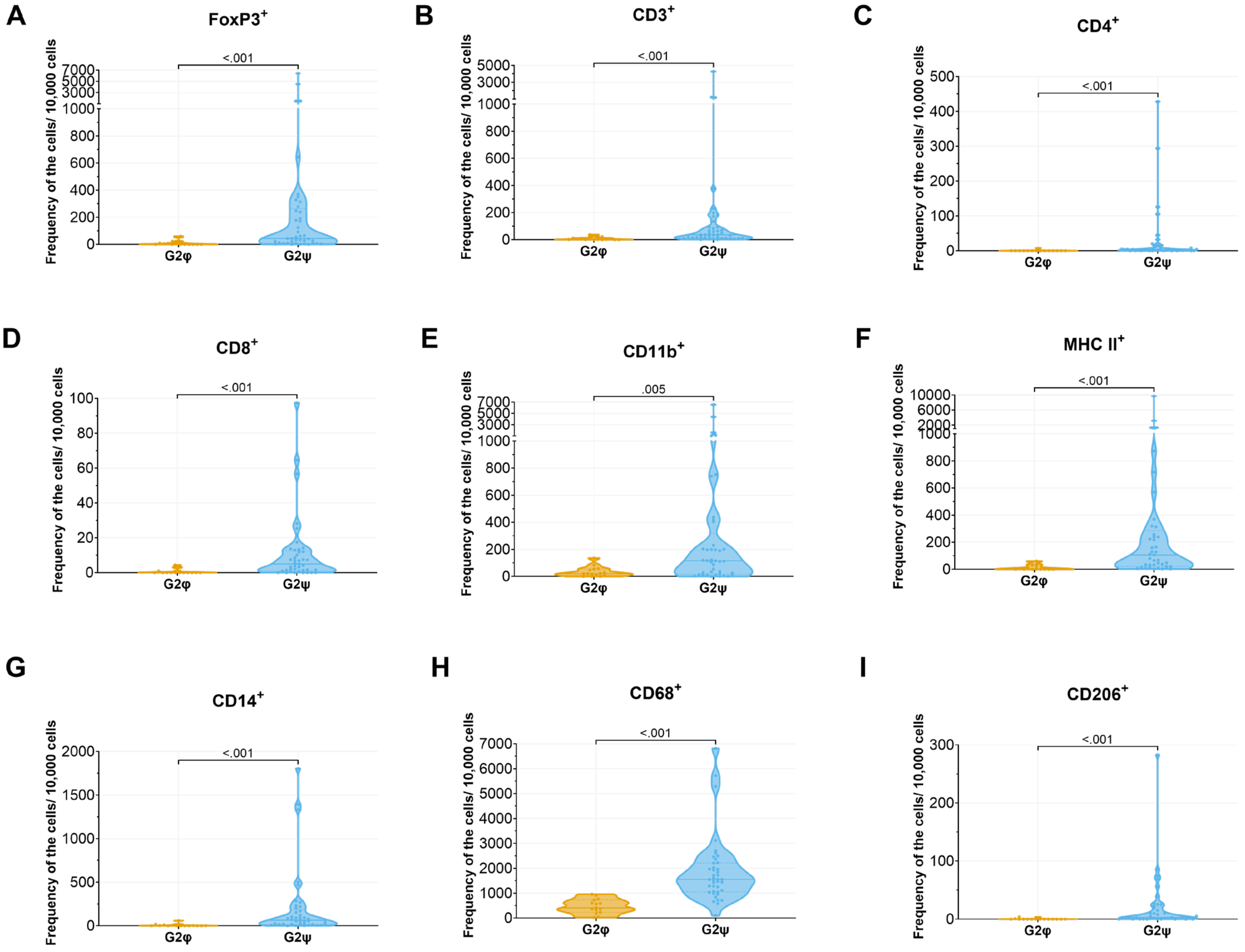


**Supplementary Figure 9:** Violin plots of primary phenotypes significant by Kruskal−Wallis testing in grade 2 cohort, stratified by subtypes.

| Cell types | Mean (SD) | | Median (IQR) | | Adjusted *P* values |
|---|---|---|---|---|---|
| | G2φ (*n* =15) | G2ψ (*n* = 41) | G2φ (*n* =15) | G2ψ (*n* = 41) | |
| All T cells ($CD3^+$, regardless of other markers in Panel 1) | 0.08 (0.1) | 2.19 (7.02) | 0.02 (0−0.11) | 0.36 (0.12 − 0.91) | **<0.001** |
| Regulatory T cells (Tregs, $CD3^+FOXP3^+$, regardless of other markers in Panel 1) | 0.02 (0.05) | 1.61 (6.07) | 0 (0−0) | 0.09 (0.02−0.44) | **<0.001** |
| T helper cells ($CD3^+CD4^+$, regardless of other markers in Panel 1) | 0 (0) | 0.1 (0.23) | 0 (0−0) | 0.01 (0−0.05) | **<0.001** |
| Cytotoxic T cells ($CD3^+CD8^+$, regardless of other markers in Panel 1) | 0.01 (0.01) | 0.1 (0.18) | 0 (0−0) | 0.04 (0.01−0.09) | **<0.001** |
| Antigen−Presenting cells (APCs, $MHCII^+CD14^-CD11b^-$ or $MHCII^+CD14^+CD11b^-$ or $MHCII^+CD14^+CD11b^+$ or $MHCII^+CD14^-CD11b^+$, regardless of other markers in Panel 2) | 0.15 (0.22) | 5.14 (15.7) | 0.02 (0−0.26) | 1.06 (0.22−2.57) | **<0.001** |
| Macrophages/microglia/monocytes ($CD14^+CD11b^-$ or $CD14^+CD11b^+$ or $CD15^+$ or $CD11b^+$, regardless of other markers in Panel 2) | 0.46 (0.44) | 7.46 (14.9) | 0.35 (0.16−0.74) | 2.23 (1.15−6.19) | **<0.001** |
| Non−M2 Macrophages/microglia/monocytes ($CD68^+CD163^-CD206^-$ or $CD68^-CD163^-CD206^+$ or $CD68^-CD163^+CD206^-$, regardless of other markers in Panel 3) | 4.71 (2.93) | 18.63 (13.26) | 4.12 (2.74−6.67) | 15.52 (10.68−21.46) | **<0.001** |
| All M2− macrophages ($CD68^+CD163^+CD206^+$ or $CD68^+CD163^-CD206^+$ or $CD68^+CD163^+CD206^-$, regardless of other markers in Panel 3) | 0 (0) | 0.17 (0.43) | 0 (0−0) | 0.03 (0.01−0.19) | **<0.001** |

**Supplementary Table 6:** Major immune cell phenotypes in grade 2 by tumor subtype.

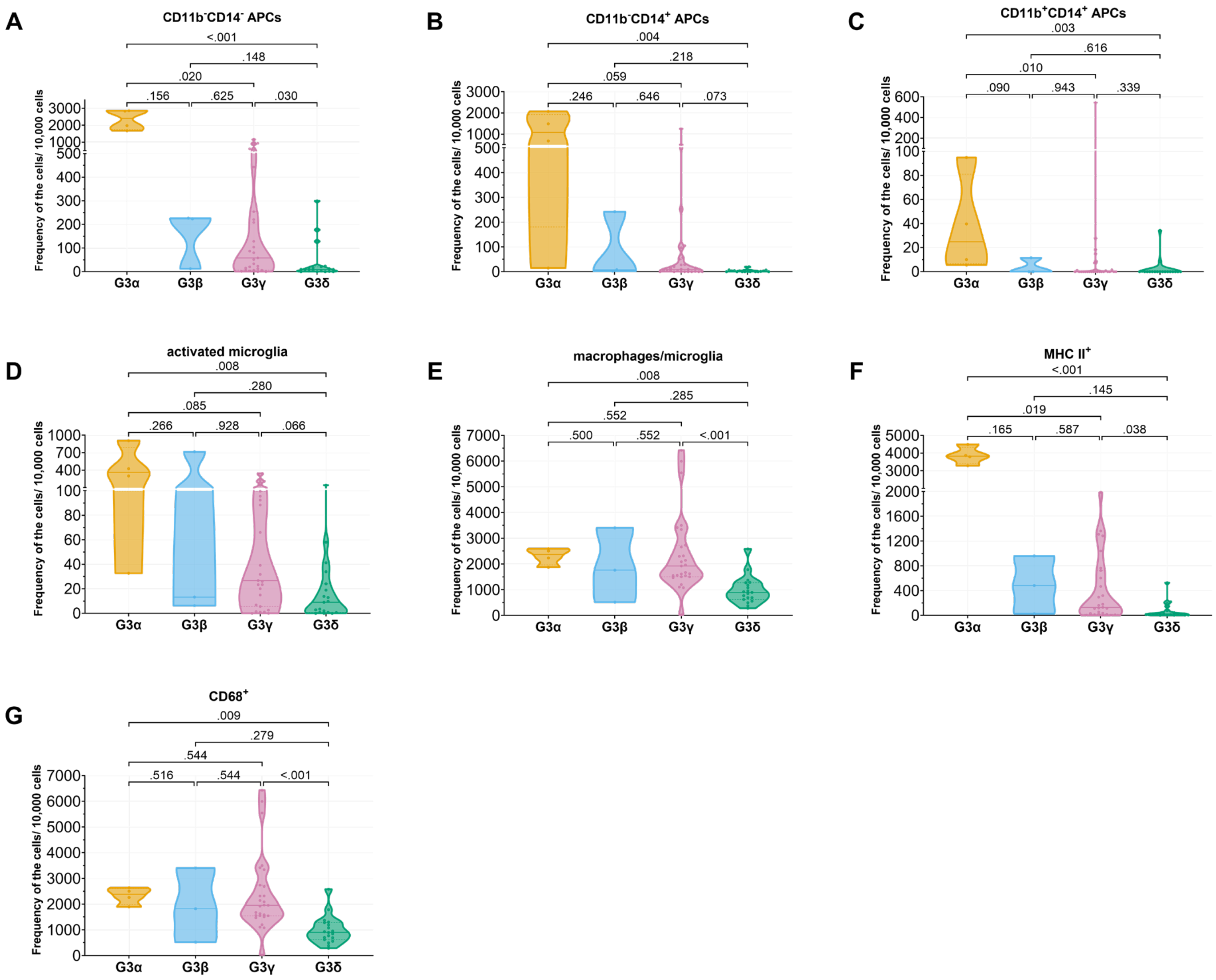


**Supplementary Figure 10:** Multi-marker secondary phenotypes and primary phenotypes that are significantly different between grade 3 subtypes when mapped into grade 4 UMAP space.

| | G3 Outliers | G3α | G3β | G3γ | G3δ | Total in row |
|---|---|---|---|---|---|---|
| **G3ε** | 1 | 0 | 0 | 0 | 10 | 11 |
| **G3ρ** | 1 | 0 | 1 | 7 | 2 | 11 |
| **G3τ** | 1 | 4 | 2 | 20 | 7 | 34 |
| **Total in column** | 3 | 4 | 3 | 27 | 19 | |

**Supplementary Table 7:** Overlaps between original grade 3 subtypes and grade 3 gliomas mapped onto the immune space of grade 4 gliomas.

| | G2ε | G2ρ | G2τ | Total in row |
|---|---|---|---|---|
| **G2φ** | 15 | 0 | 0 | 15 |
| **G2ψ** | 7 | 10 | 24 | 41 |
| **Total in column** | 22 | 10 | 24 | |

**Supplementary Table 8:** Overlaps between original grade 2 subtypes and grade 2 gliomas mapped onto the immune space of grade 3 gliomas.

**Supplementary Note 1: Survival Analysis Based on Primary and Multi−marker Secondary Phenotypes.**

After scaling multi−marker secondary phenotypes by z−score normalization and removing sparse variables, correlation filtering was performed to eliminate phenotypes that could cause high multicollinear issues. Multicollinearity was assessed using the condition number, with a threshold of 100. To ensure robustness to variable ordering, 10000 random permutations of phenotype order were evaluated, sequentially adding variables to the feature set while maintaining the condition number below the predefined threshold.

For patients with grade 4 gliomas, to specify multi−marker secondary phenotypes, we used the R package "glmnet" (v4.1.9) and trained elastic net penalized cox regression models with ten−fold cross−validation (alpha = 0.5, balancing L1 and L2 regularization) to promote stability in variable selection[1,2]. For patients with grade 3 gliomas, five−fold cross−validation was employed due to the limited number of patients and events (death), with the model complexity guided by the available events per variable (EPV)[3]. Additionally, the elastic net procedure was repeated 1000 times with different random seeds to ensure robustness of variable selection.

**Supplementary Note 2: Details of cox regression models.**

For the grade 4 cohort, prior to elastic net regularization, MHC $II^{-}$ monocytes were excluded because they violated the proportional hazards assumption and were consistently retained by elastic net. This multi-marker secondary phenotype was evaluated separately using univariable Cox proportional hazards regression, with MHC $II^{-}$ monocytes associated with unsignificant increased risk (HR = 1.09, 95% CI: 0.96−1.25, p = 0.173). In addition, double-negative T cells and cytotoxic T cells were removed due to substantial multicollinearity. This decision was further supported by 10,000 random permutations of phenotype ordering, under which a single backward-selection procedure repeatedly identified these two variables for exclusion. Furthermore, bidirectional stepwise selection applied to the elastic net selected model did not remove any feature.

For the grade 3 cohort, features with extreme sparsity (≤10% nonzero observations), including $PD1^{+}CD3^{-}$ cells, $PD1^{+}$T lymphocytes, and myeloid-like cells, were excluded. And $CD11b^{+}CD14^{+}$ APCs, T helper cells, cytotoxic T cells, innate−like T cells, Tregs, double−positive T cells were removed due to multicollinearity before elastic net regularization. To mitigate the risk of overfitting caused by the limited numbers of observed events in the grade 3 cohort, we applied elastic net penalized cox regression with five−fold cross validation, with the model complexity guided by the 10 available events per variable. Following 1,000 repeated runs with randomly seeds, only three multi−marker secondary phenotypes were consistently selected and retained in the final model.

For the grade 2 cohort, due to the limited number of events (n = 10), only univariable cox proportional hazards models were performed. Prior to analysis, multi−marker secondary phenotypes with extreme sparsity (≤10% nonzero observations), including double−positive T cells, PD1+ T lymphocytes, PD1+CD3− cells, myeloid-like cells, M2 macrophages, B cells and CD8+ Tregs were excluded.

As no significant difference in OS was observed between IDH1-wt grade 4 tumors and other grade 4 tumors by the Mann−Whitney U test (P = 0.22), results from all grades 4 tumors are presented together in this survival analysis.

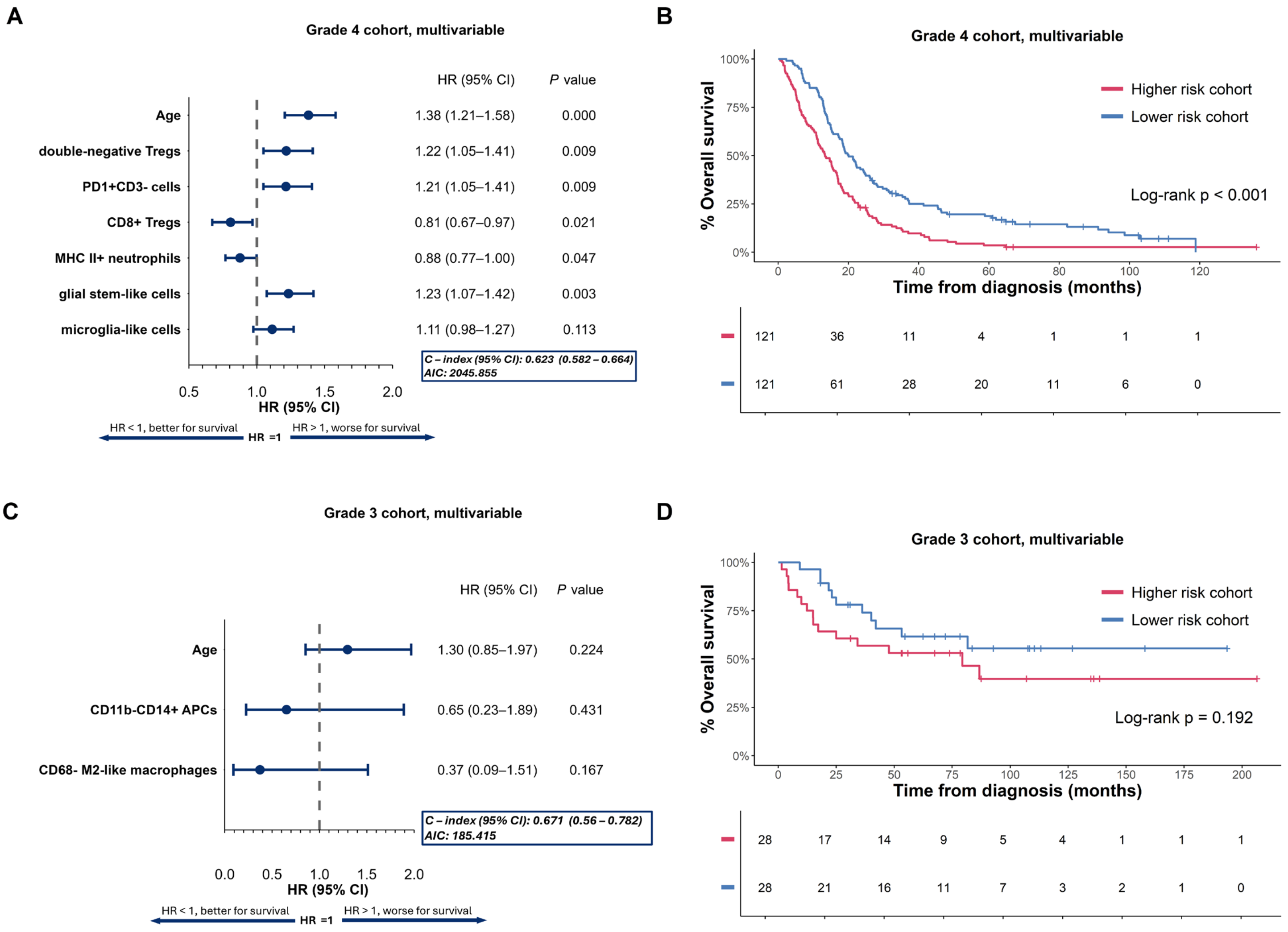


**Supplementary Figure 11.** Multivariable cox models of multi−marker secondary phenotypes, and risk score prediction for OS in grade 4 and grade 3 glioma. **(A)** & **(C)**: Forest plots multivariable cox proportional hazards models for OS, evaluating multi−marker secondary phenotypes in grade 4 and grade 3 patient cohorts. Hazard ratios (HR) and 95% confidence intervals (95% CI) are shown for each selected variable. P values are tested by two−sided Wald test. HR > 1 indicates worse survival, while HR < 1 indicates better survival. **(B)** & **(D)**: Corresponding Kaplan−Meier survival curves for OS based on risk scores predicted by the multivariable cox models for grade 4 and grade 3 patient cohorts. Patients were classified into higher−risk and lower−risk groups using the median risk score as the cutoff. Log−rank p values were calculated using two−sided log−rank tests.

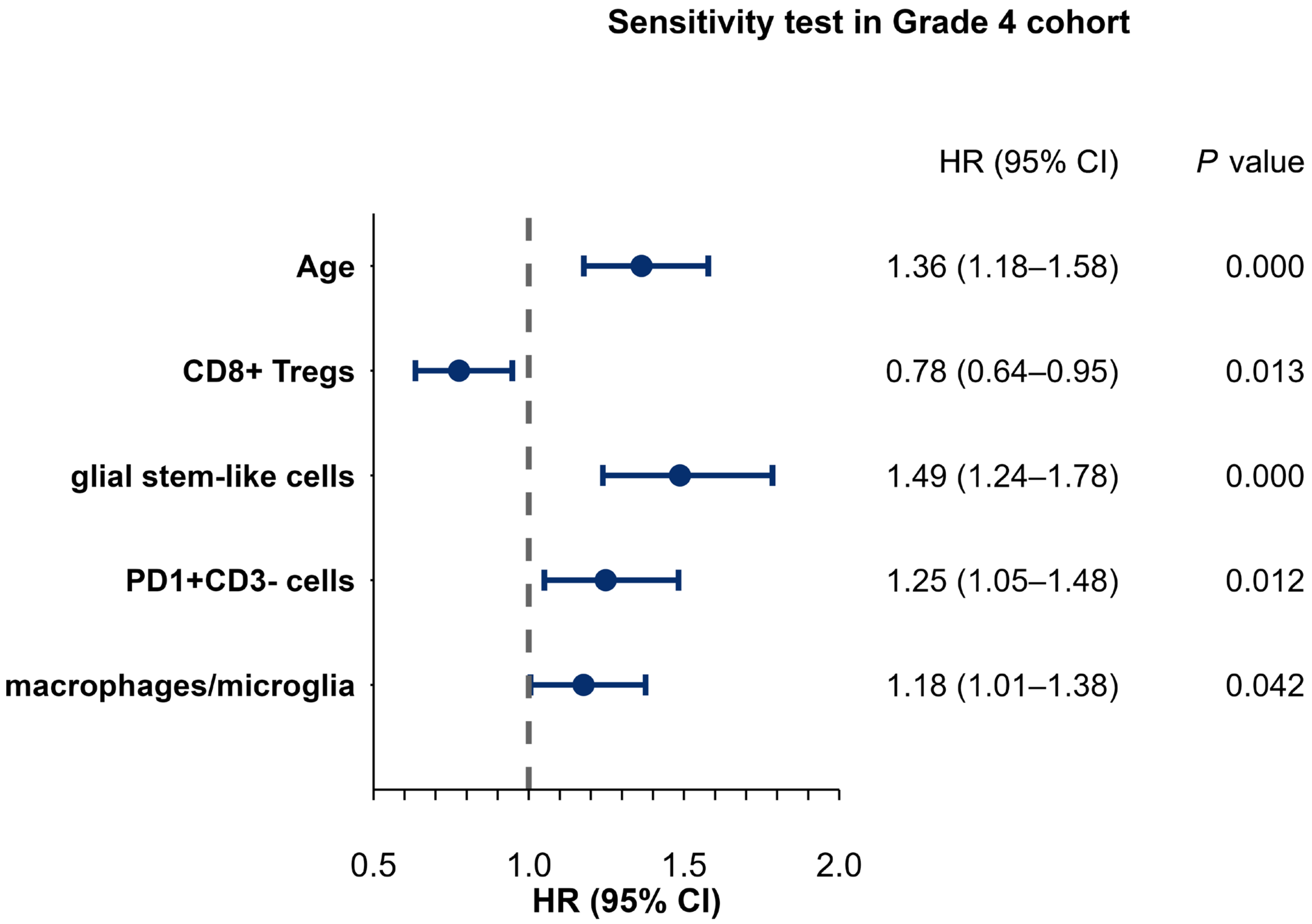


**Supplementary Figure 12:** Sensitivity analysis in the grade 4 cohort restricting to *IDH1* wildtype tumors.

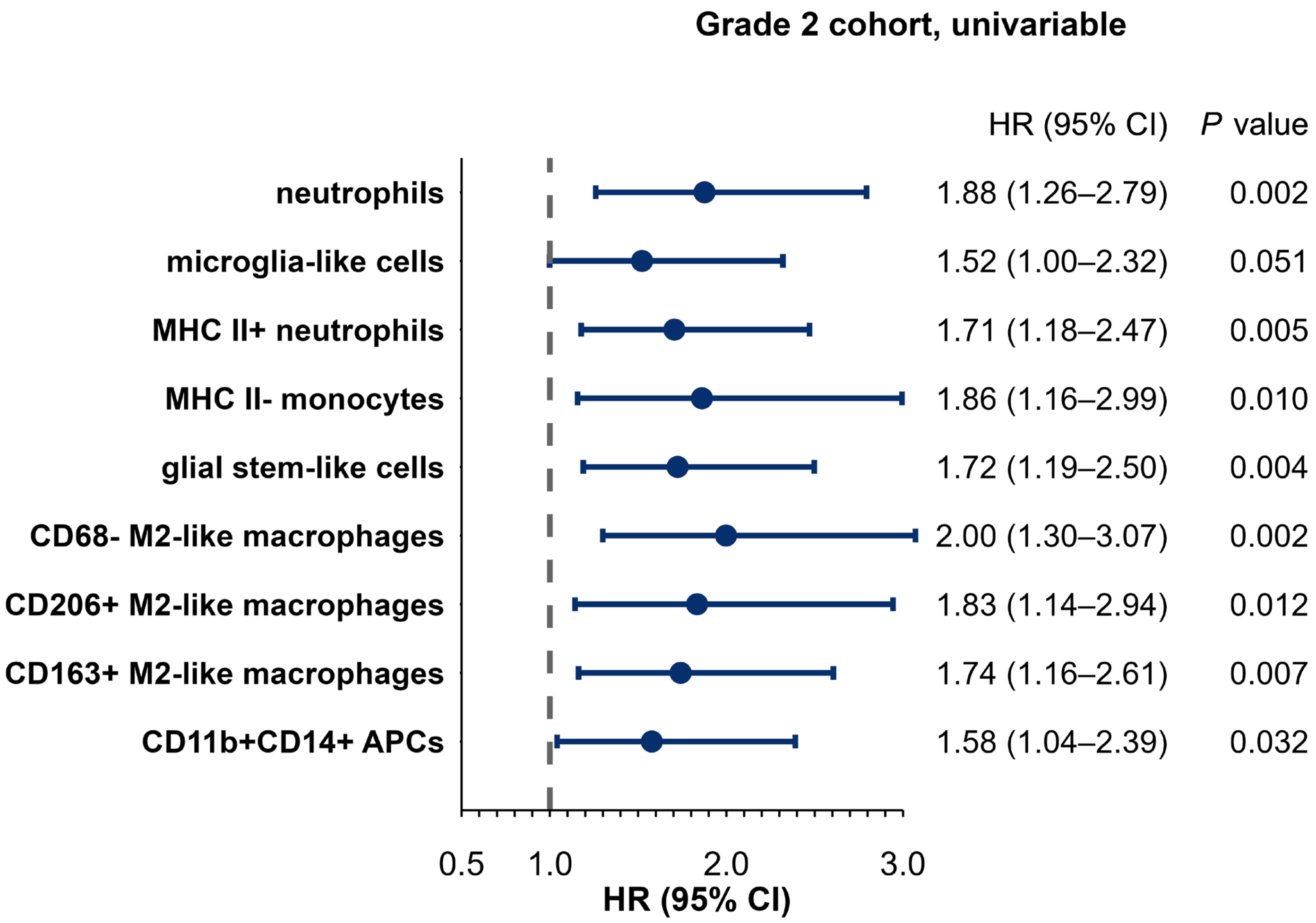


**Supplementary Figure 13:** Forest plots univariable cox proportional hazards models for OS, evaluating multi−marker secondary phenotypes in grade 2 patient cohorts. Hazard ratios (HR) and 95% confidence intervals (95% CI) are shown for variables present (non-zero) in at least 10% of samples. P values are tested by two−sided Wald test. HR > 1 indicates worse survival, while HR < 1 indicates better survival.

**Supplementary Note 3: Comparison of Different Grades**

To explore differences in primary phenotypes and multi−marker secondary phenotypes across different grades, Kruskal–Wallis tests, followed by Dunn's test with FDR correction, were performed and results are visualized by violin plots.

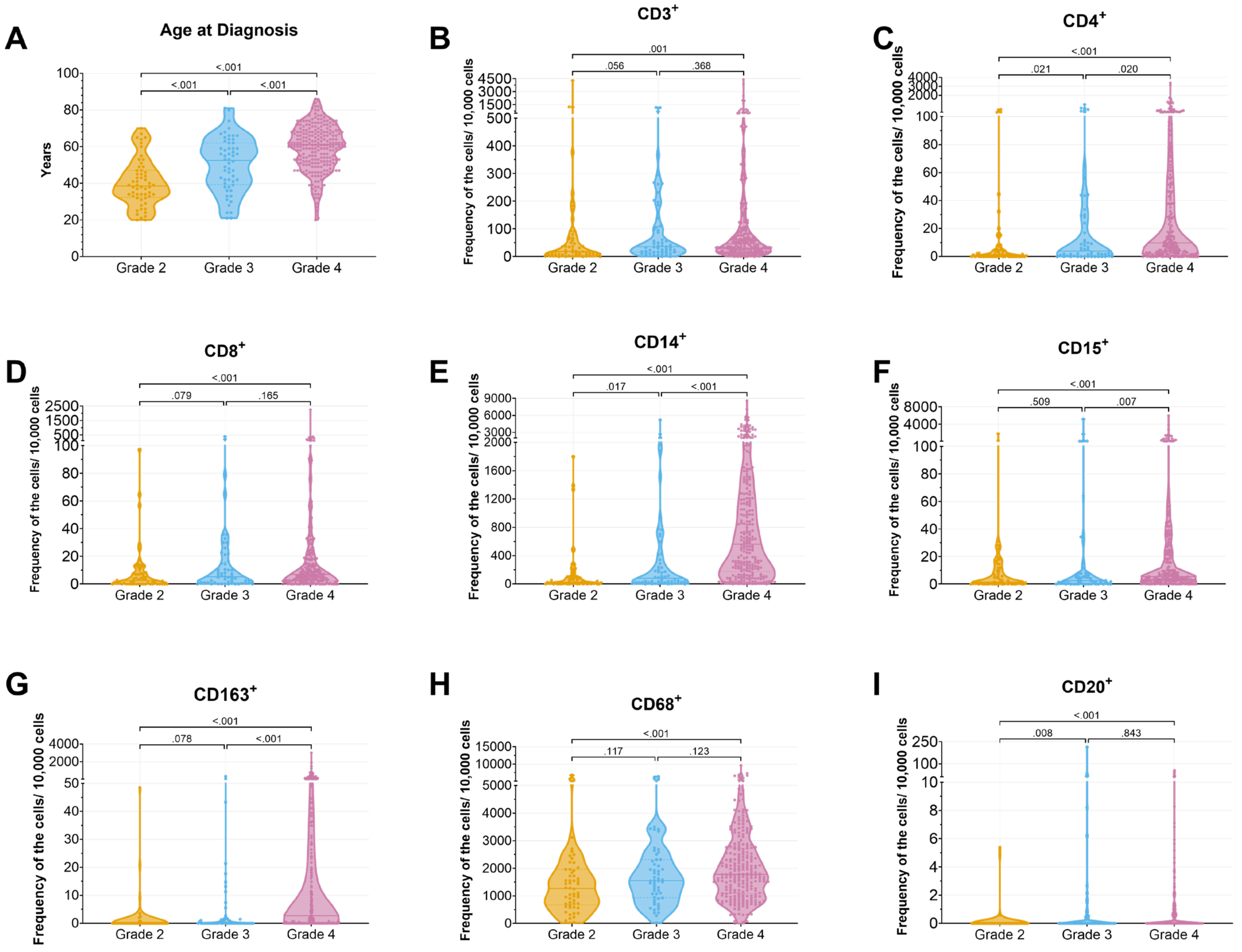


**Supplementary Figure 14:** Violin plots of age and all significant primary phenotypes by Kruskal−Wallis testing in comparison different grades. Pairwise comparisons between grades were performed using Dunn's multiple comparison test (p values were adjusted by FDR). Solid lines indicate median values; dashed lines represent the 25th and 75th percentiles.

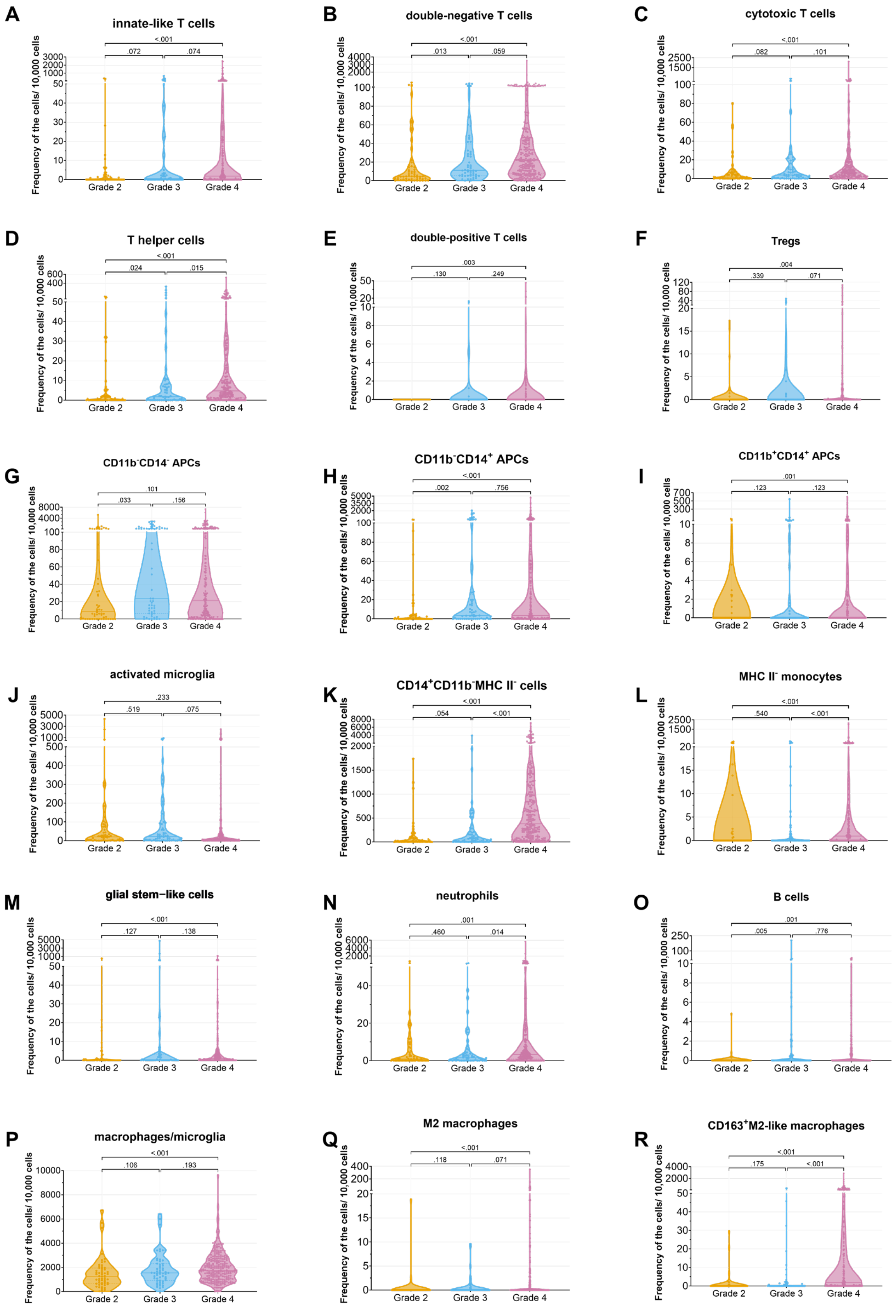


**Supplementary Figure 15:** Violin plots of all significant multi-marker secondary phenotypes by Kruskal−Wallis testing in comparison different grades. Pairwise comparisons between grades were performed using Dunn's multiple comparison test (p values were adjusted by FDR). Solid lines indicate median values; dashed lines represent the 25th and 75th percentiles.

| Cell types | Mean (SD) | | | Median (IQR) | | | Adjusted *P* values |
|---|---|---|---|---|---|---|---|
| | Grade 2 (n = 56) | Grade 3 (n = 56) | Grade 4 (n = 242) | Grade 2 (n = 56) | Grade 3 (n = 56) | Grade 4 (n = 242) | |
| All T cells ($CD3^+$, regardless of other markers in Panel 1) | 1.63 (6.06) | 1.22 (2.34) | 1.73 (4.68) | 0.18 (0.07–0.65) | 0.35 (0.16–1.06) | 0.46 (0.19–1.22) | **0.002** |
| Regulatory T cells (Tregs, $CD3^+FOXP3^+$, regardless of other markers in Panel 1) | 1.19 (5.23) | 0. 36 (0.78) | 0.52 (2.35) | 0.06 (0. 00–0. 21) | 0.05 (0. 00–0.21) | 0.03 (0. 00–0.14) | 0.523 |
| T helper cells ($CD3^+CD4^+$, regardless of other markers in Panel 1) | 0.07 (0.20) | 0. 25 (0.72) | 0.27 (0.62) | 0.00 (0.00–0.02) | 0.02 (0.00–0.08) | 0.05 (0.01–0.23) | **< 0.001** |
| Cytotoxic T cells ($CD3^+CD8^+$, regardless of other markers in Panel 1) | 0.07 (0. 16) | 0. 18 (0.52) | 0.29 (1.45) | 0.02 (0.00–0.06) | 0.04 (0.01–0.15) | 0.06 (0.02–0.18) | **< 0.001** |
| Antigen−Presenting cells (APCs, $MHCII^+CD14^-CD11b^-$ or $MHCII^+CD14^+CD11b^-$ or $MHCII^+CD14^+CD11b^+$ or $MHCII^+CD14^-CD11b^+$, regardless of other markers in Panel 2) | 3.80 (13.57) | 5.80 (10.76) | 5.97 (13.04) | 0.42 (0.06–2.19) | 1.08 (0.20–5.45) | 0.58 (0.11–4.22) | 0.241 |
| Macrophages/microglia/monocytes ($CD14^+CD11b^-$ or $CD14^+CD11b^+$ or $CD15^+$ or $CD11b^+$, regardless of other markers in Panel 2) | 5.59 (13.09) | 7.80 (12.67) | 15.60 (21.47) | 1.29 (0.41–3.52) | 1.19 (0.66–9.46) | 8.36 (2.88–19.04) | **< 0.001** |
| Non−M2 Macrophages/microglia/monocytes ($CD68^+CD163^-CD206^-$ or $CD68^-CD163^-CD206^+$ or $CD68^-CD163^+CD206^-$, regardless of other markers in Panel 3) | 14.90 (12.99) | 18.35 (13.23) | 19.94 (12.60) | 12.65 (6.97–18.98) | 15.45 (9.43–23.03) | 17.46 (10.66–26.16) | **0.002** |
| All M2− macrophages ($CD68^+CD163^+CD206^+$ or $CD68^+CD163^-CD206^+$ or $CD68^+CD163^+CD206^-$, regardless of other markers in Panel 3) | 0.13 (0. 38) | 0.14 (0.47) | 1.13 (3.16) | 0.02 (0.00–0.11) | 0.01 (0.00–0.09) | 0.08 (0.00–0.66) | **< 0.001** |

**Supplementary Table 9:** Summary of features by glioma grade. Mean (standard deviation, SD) and median (interquartile range, IQR, Q1−Q3) of percentage of selected cells across glioma groups: grade 2, grade 3 and grade 4. Overall group differences were assessed by the Kruskal–Wallis test. P values are adjusted by FDR. Significant p−values are shown in bold ($p < 0.05$).

| Cell types | Mean (SD) | | | Median (IQR) | | | Adjusted *P* values |
|---|---|---|---|---|---|---|---|
| | Mutation (n = 19) | Wildtype (n = 206) | Not done (n = 17) | Mutation (n = 19) | Wildtype (n = 206) | Not done (n = 17) | |
| At least one of (FOXP3, CD3, CD4, CD8 in Panel 1) expressed positive | 1.26 (1.53) | 4.37 (9.57) | 2.59 (3.25) | 0.48 (0.30−1.87) | 0.94 (0.38–3.03) | 1.36 (0.46−3.96) | 0.33 |
| At least one of (CD11b, MHC II, CD14, CD15, CD33 in Panel 2) expressed positive | 11.49 (10.55) | 16.86 (19.04) | 26.86 (26.39) | 8.74 (1.28–17.45) | 9.63 (3.69–23.04) | 15.02 (2.44−48.45) | 0.33 |
| At least one of (CD163, CD68, CD20, CD19, CD206 in Panel 3) expressed positive | 18.11 (9.66) | 21.15 (14.37) | 23.58 (13.08) | 16.74 (10.7−21.78) | 17.73 (11.01–27.4) | 23.62 (17.4−32.91) | 0.33 |

**Supplementary Table 10:** Summary of features in grade 4 cohort by *IDH1* status.
